# Associations and prognostic accuracy of electrolyte imbalances in predicting poor COVID-19 outcome: a systematic review and meta-analysis

**DOI:** 10.1101/2021.11.19.21266563

**Authors:** Harris Jun Jie Muhammad Danial Song, Alys Zhi Qin Chia, Benjamin Kye Jyn Tan, Chong Boon Teo, Horng Ruey Chua, Miny Samuel, Adrian Kee

**Affiliations:** Yong Loo Lin School of Medicine, National University of Singapore (NUS), Singapore; Department of Medicine, National University Hospital (NUH), Singapore; Research Support Unit, Yong Loo Lin School of Medicine, National University of Singapore (NUS), Singapore

**Keywords:** Electrolytes, Severe Acute Respiratory Syndrome, Hypernatremia, Death Risk, Intensive Care, Respiratory Medicine

## Abstract

**Background:** Serum electrolyte imbalances are highly prevalent in COVID-19 patients. However, their associations with COVID-19 outcomes are inconsistent, and of unknown prognostic value.

**Objectives:** To systematically clarify the associations and prognostic accuracy of electrolyte imbalances (sodium, calcium, potassium, magnesium, chloride and phosphate) in predicting poor COVID-19 clinical outcome.

**Methods:** PubMed, Embase and Cochrane Library were searched. Odds of poor clinical outcome (a composite of mortality, intensive-care unit (ICU) admission, need for respiratory support and acute respiratory distress syndrome) were pooled using mixed-effects models. The associated prognostic sensitivity, positive and negative likelihood ratios (LR+, LR-) and predictive values (PPV, NPV; assuming 25% pre-test probability), and area under the curve (AUC) were computed.

**Results:** We included 28 observational studies from 953 records with low to moderate risk-of-bias. Hyponatremia (OR=2.08, 95%CI=1.48-2.94, I^2^=93%, N=8), hypernatremia (OR=4.32, 95%CI=3.17-5.88, I^2^=45%, N=7) and hypocalcemia (OR=3.31, 95%CI=2.24-4.88, I^2^=25%, N=6) were associated with poor COVID-19 outcome. These associations remained significant on adjustment for covariates such as demographics and comorbidities. Hypernatremia was 97% specific in predicting poor outcome (LR+ 4.0, PPV=55%, AUC=0.80) despite no differences in CRP and IL-6 levels between hypernatremic and normonatremic patients. Hypocalcemia was 76% sensitive in predicting poor outcome (LR- 0.44, NPV=87%, AUC=0.71). Overall quality of evidence ranged from very low to moderate.

**Conclusion:** Hyponatremia, hypernatremia and hypocalcemia are associated with poor COVID-19 clinical outcome. Hypernatremia is 97% specific for a poor outcome and the association is independent of inflammatory marker levels. Further studies should evaluate if correcting these imbalances help improve clinical outcome.

## INTRODUCTION

Since the first case of the coronavirus disease 2019 (COVID-19) in December 2019,[1] more than 200 million people have been diagnosed and cumulative deaths have exceeded 4.25 million as of 4^th^ August 2021.[2] As majority of cases develop mild symptoms or are asymptomatic,[3, 4] many have identified several risk factors associated with poor outcomes to better triage and allocate scarce health resources. [5–11]

Recently, multiple observational studies reported the prevalence of electrolyte disorders amongst COVID-19 patients and have suggested its association with COVID-19 severity. [12, 13] For example, Tzoulis et al. found that adults with COVID-19 and dysnatremia was associated with a higher risk for mechanical ventilation and mortality in adults.[14] Some have postulated that COVID-19 using the cell entry receptor angiotensin-converting enzyme 2 (ACE2) could result in fluid and electrolyte imbalances as it is also one of the key enzymes in the renin-angiotensin system (RAS).[15, 16] Since serum electrolytes are one of the most basic laboratory tests readily performed on a regular basis inpatient,[17] they may serve as additional tools in the triaging of resources.

While previous meta-analyses have reported associations of hypocalcemia and hyponatremia with COVID-19 severity,[18, 19] recent published studies have also suggested associations of other electrolyte imbalances such as dysnatremia, dyskalemia, dysmagnesemia and dyschloremia with COVID-19 severity.[14, 20–24] However, the association of these electrolyte imbalances with COVID-19 outcomes are inconsistent, and their pooled prognostic value are not yet determined. Furthermore, the association with acute kidney injury (AKI) and acute respiratory distress syndrome (ARDS) were not included despite them being highly incident in hospitalized COVID-19 patients.[25, 26] Thus, we sought to conduct a systematic review and meta-analysis to investigate the association of electrolyte imbalances with COVID-19 outcomes. Given the disruption and economic burden the pandemic has brought to countries and our daily lives,[27, 28] coupled with the immense toll on some healthcare systems,[29] this review is both timely and clinically relevant to help improve risk stratification and resource allocation.

## METHODS

This review is registered on PROSPERO (<u>CRD42021257711</u>) and reported according to Preferred Reporting Items for Systematic Reviews and Meta-analysis (PRISMA) guidelines (**Supplementary Table S1**).[30]

### Search Strategy

We searched three databases (PubMed, Embase and Cochrane Library) from inception till 22nd May 2021 using search terms related to COVID-19 and electrolyte imbalances concerning the electrolytes sodium, calcium, potassium, magnesium, chloride and phosphate (**Supplemental Methods**). We also hand-searched the bibliography of included articles and relevant reviews but included no additional studies.

### Study Selection, Data Extraction, Risk of Bias Assessment and Quality of Evidence

Two authors independently selected relevant studies, extracted key data and assessed risk of bias in a blinded manner using the online platform Rayyan.[31] We accepted observational studies published as full-length articles in peer-reviewed journals that reported the associations of electrolyte imbalances in patients diagnosed with COVID-19 that were either higher (e.g. hypernatremia) or lower (e.g. hyponatremia) than the normal physiological range. Outcomes of interest included mortality, intensive care unit (ICU) admissions, respiratory support, acute respiratory distress syndrome (ARDS) and/or acute kidney injury (AKI). We also included articles that reported the laboratory parameters of serum creatinine (SCr), C-reactive protein (CRP) and interleukin-6 (IL-6) at admission. We excluded case reports, reviews and letters, as well as articles published in languages other than English. We extracted key data and assessed the risk of bias using the Newcastle-Ottawa Scale (**Supplemental Methods).** Overall quality of evidence was assessed using the GRADE framework.[32]

### Statistical Analyses

We did separate meta-analyses for each type of electrolyte imbalance to compute a summary estimate of the association of the electrolyte imbalance with the above specified clinical outcomes using an inverse variance-weighted mixed-effects model (**Supplemental Methods)**. We pooled odds ratios for dichotomous outcomes and mean differences for continuous outcomes including laboratory parameters and hospitalization time (**Supplemental Methods)**. We defined poor outcome as a composite of mortality, ICU admission, respiratory support (oxygen supplementation, invasive and/or non-invasive ventilation) and ARDS due to their resource-intensive nature, in line with previous landmark studies on severe COVID.[33, 34] We generated a summary receiver operator characteristic curve (SROC), Fagan’s nomogram, coupled funnel plots and calculated the area under the curves (AUC), sensitivity, specificity, positive (LR+) and negative likelihood ratios (LR-), and positive (PPV) and negative predictive values (NPV) to evaluate the performance and prognostic value of each type of electrolyte imbalance in predicting the unadjusted odds of poor outcome. We assessed and considered between-study heterogeneity as significant if the I^2^ statistic was ≥50% and the p-value for the Q-test was <0.10.[35] To investigate potential sources of heterogeneity, we pre-specified various study-level characteristics (**Supplemental Methods**) to perform subgroup or sensitivity analyses. We conducted all analyses using RevMan (version 5.4), Stata (version 17) and RStudio (version 1.4) using the *meta* package (version 4.18).

## RESULTS

We screened 953 records after removing duplicates and subsequently identified 28 studies for inclusion after screening based on the title and abstract, followed by screening based on full-text (**Figure 1**).[14, 20–24, 36–57] Twenty-four studies were included in our various meta-analyses.[14, 20–23, 36–43, 45, 47–56]

**Figure 1:**
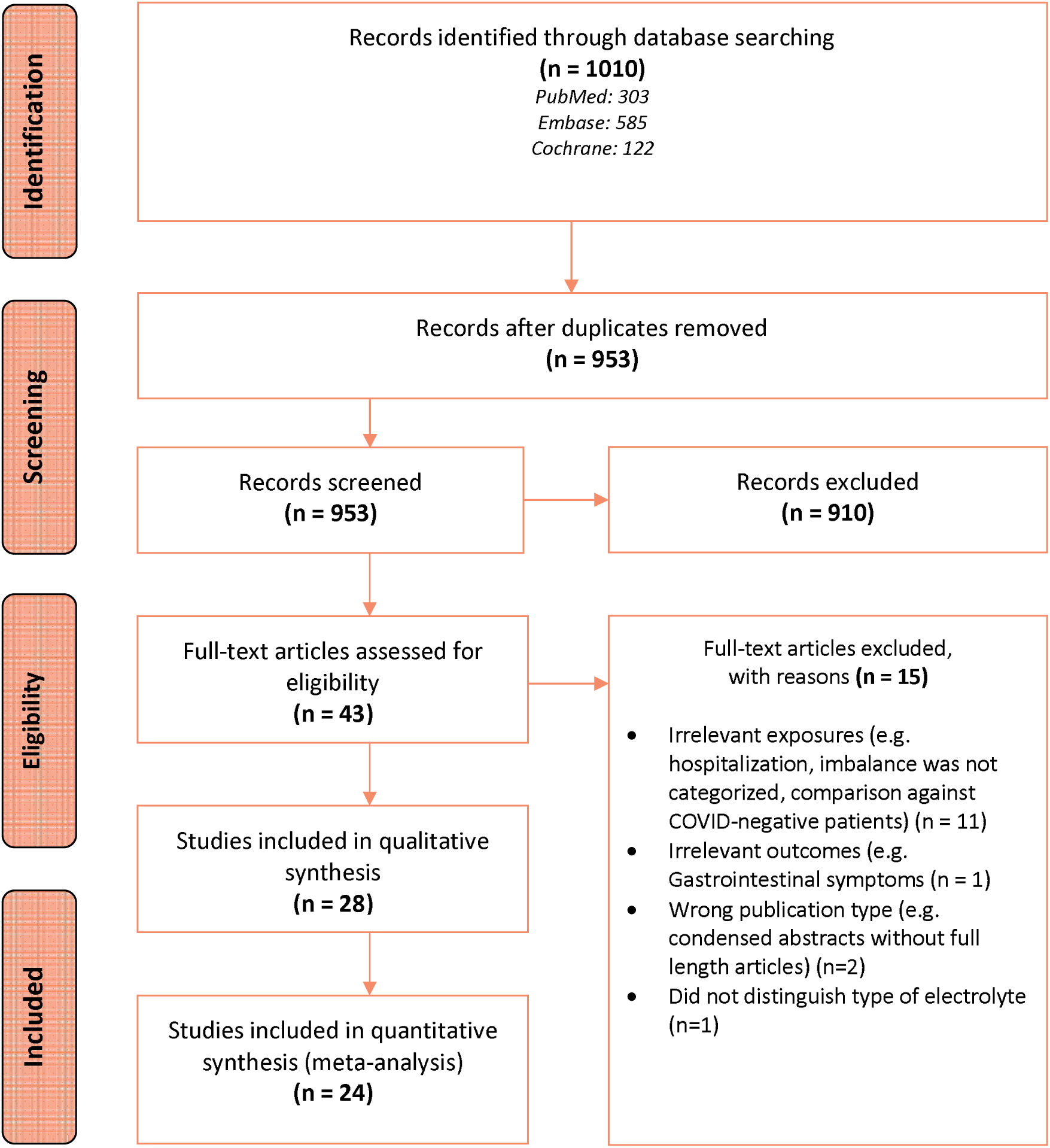
PRISMA flow diagram of the study selection process.

### Study Characteristics

Of the 28 included studies (**Table 1**), 21 were retrospective cohorts,[14, 20, 21, 23, 24, 39–47, 49, 51, 53–57] four were prospective cohorts,[22, 36–38] two were cross-sectional[48, 52] and one was a case-control study.[50] Sensitivity analyses excluding non-cohort studies did not change our findings substantially. A total of 14, 10, two studies and one study were conducted in Asia,[24, 36, 40, 43–46, 48–50, 52, 53, 56, 57] Europe,[14, 20, 22, 23, 37, 39, 41, 47, 54, 55] America[21, 42] and Africa[38] respectively. One study spanned across America, Asia and Europe.[51] When assessed using the Newcastle-Ottawa Scale, twenty and eight studies had a moderate and low risk of bias respectively. Overall quality of evidence ranged from very low to moderate.

**Table 1:**
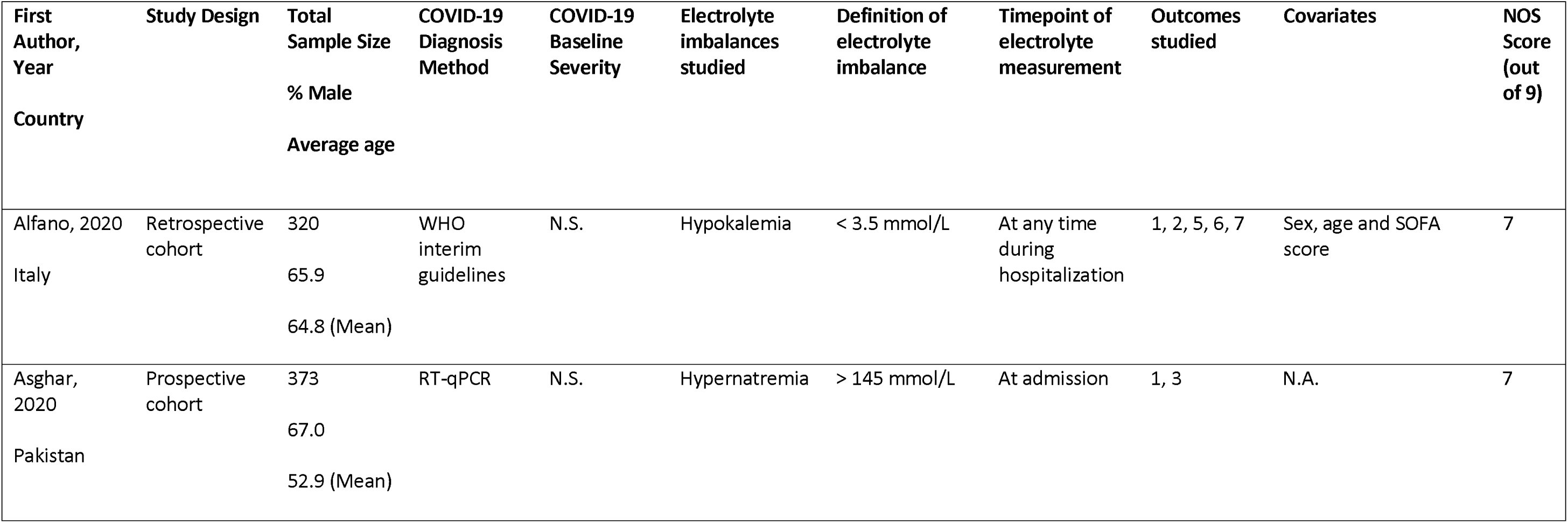
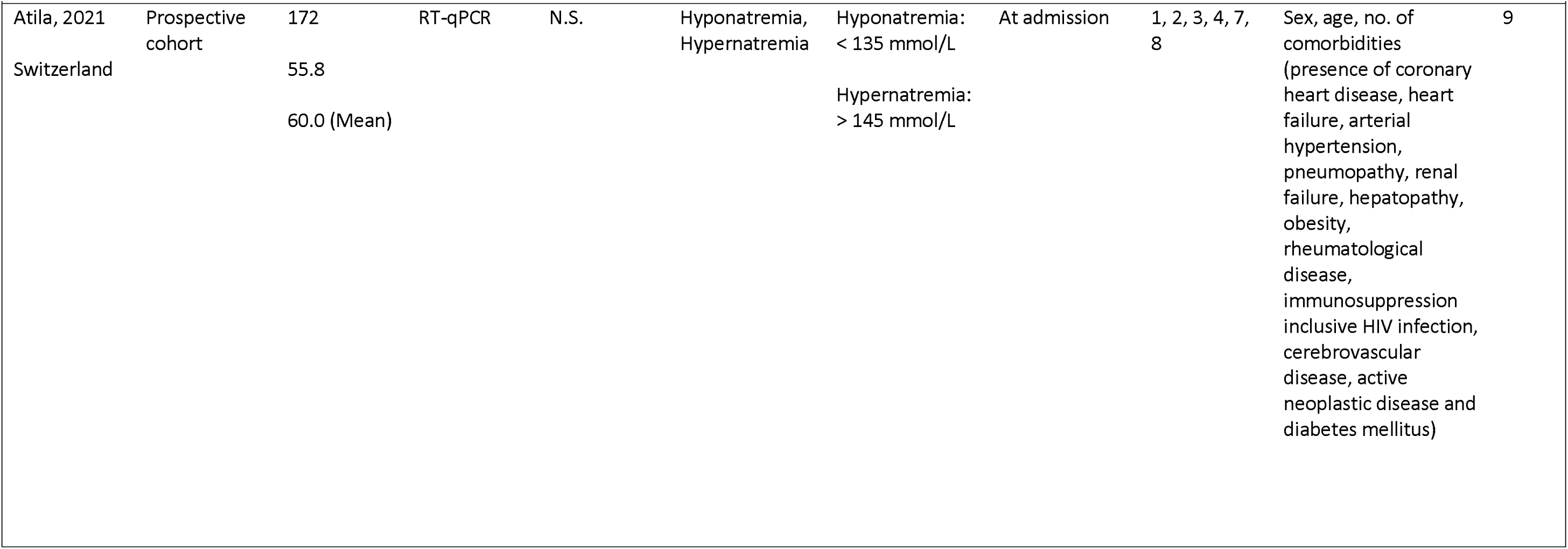
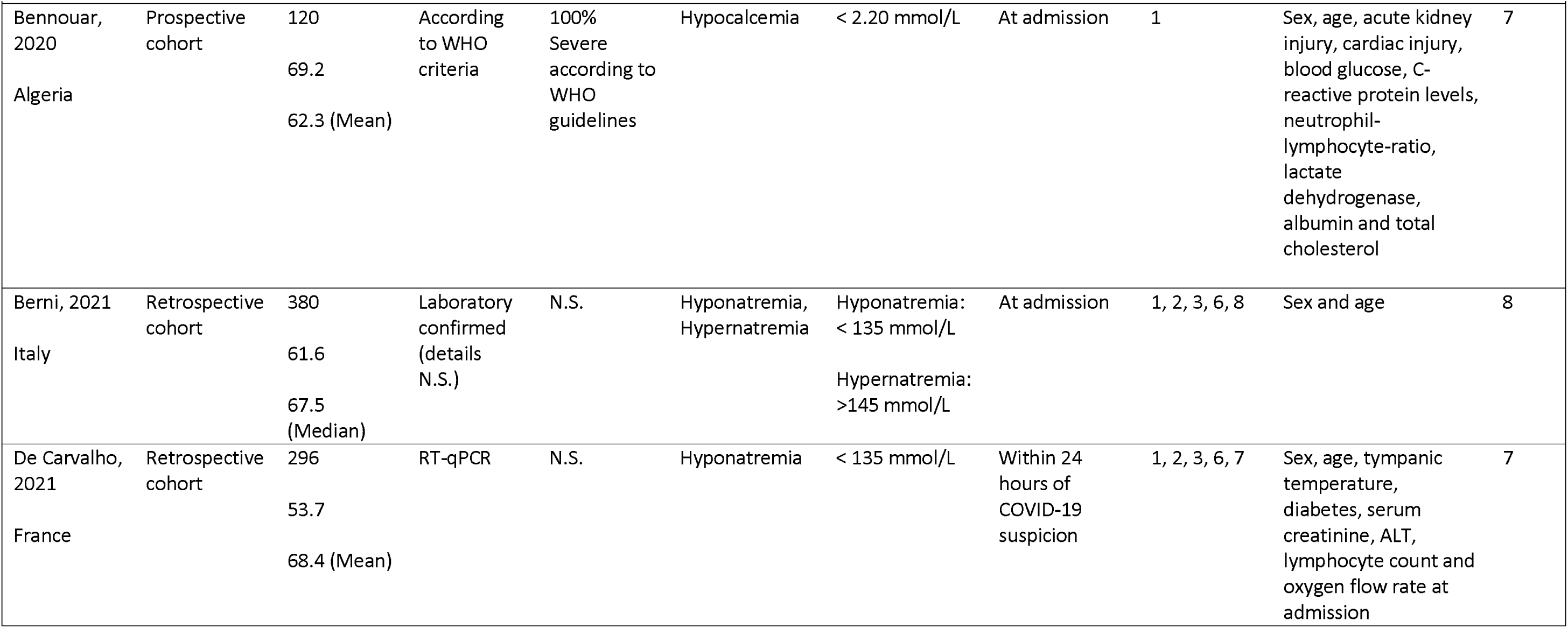
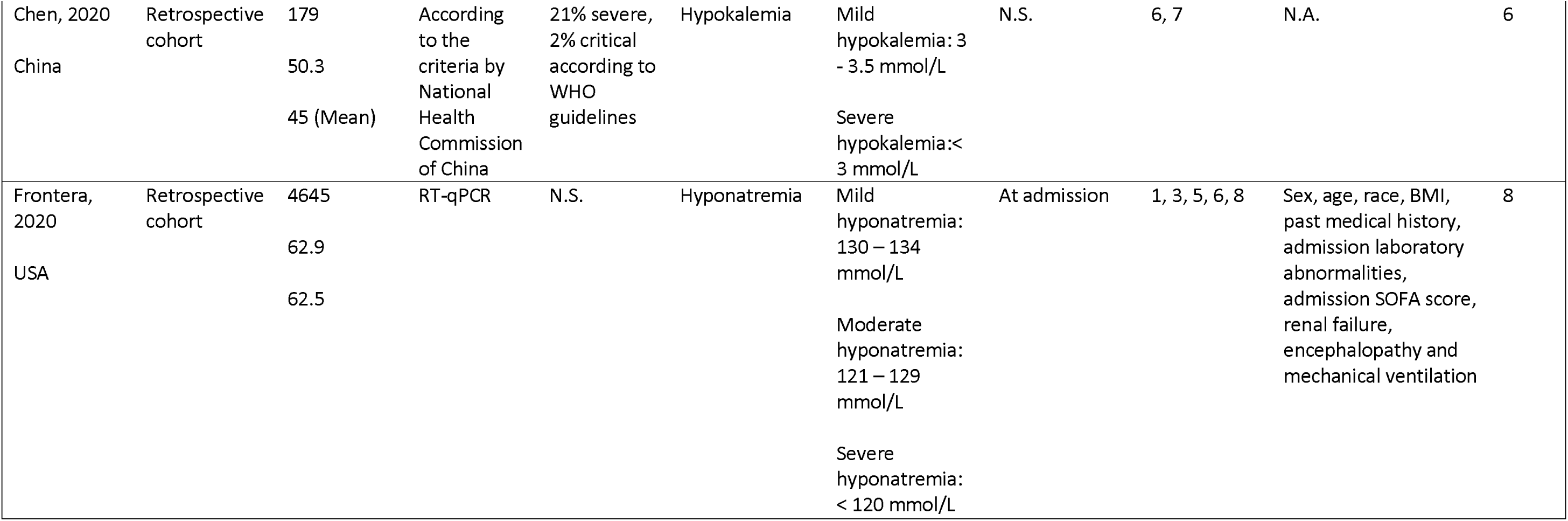
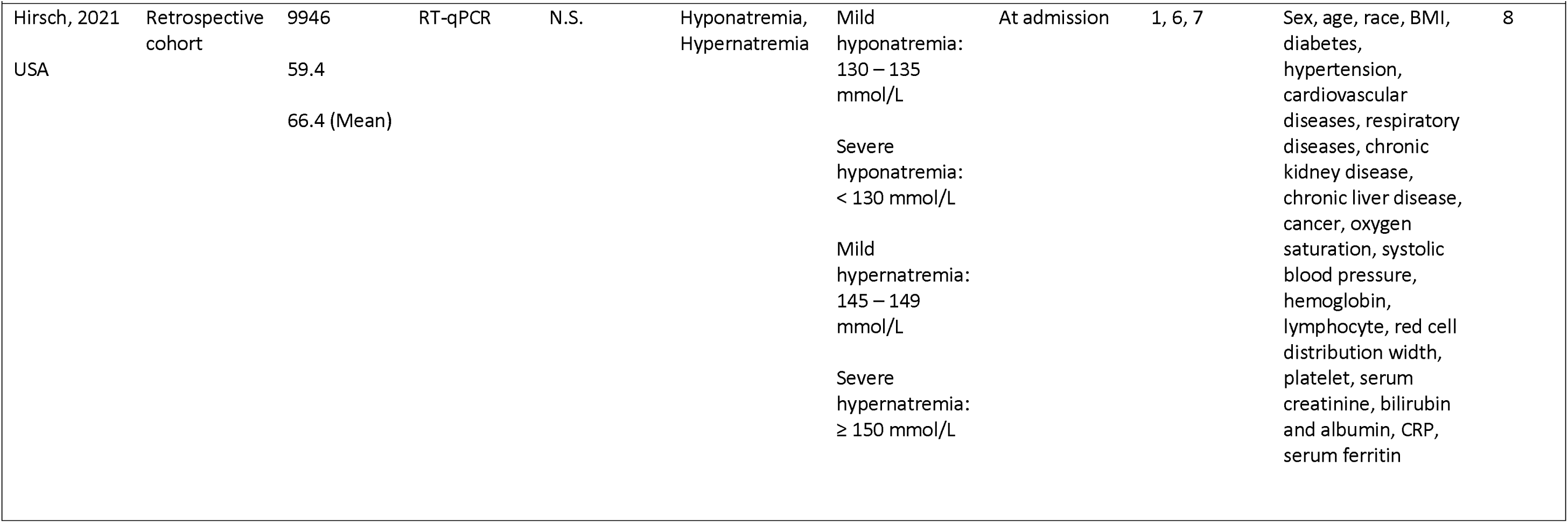
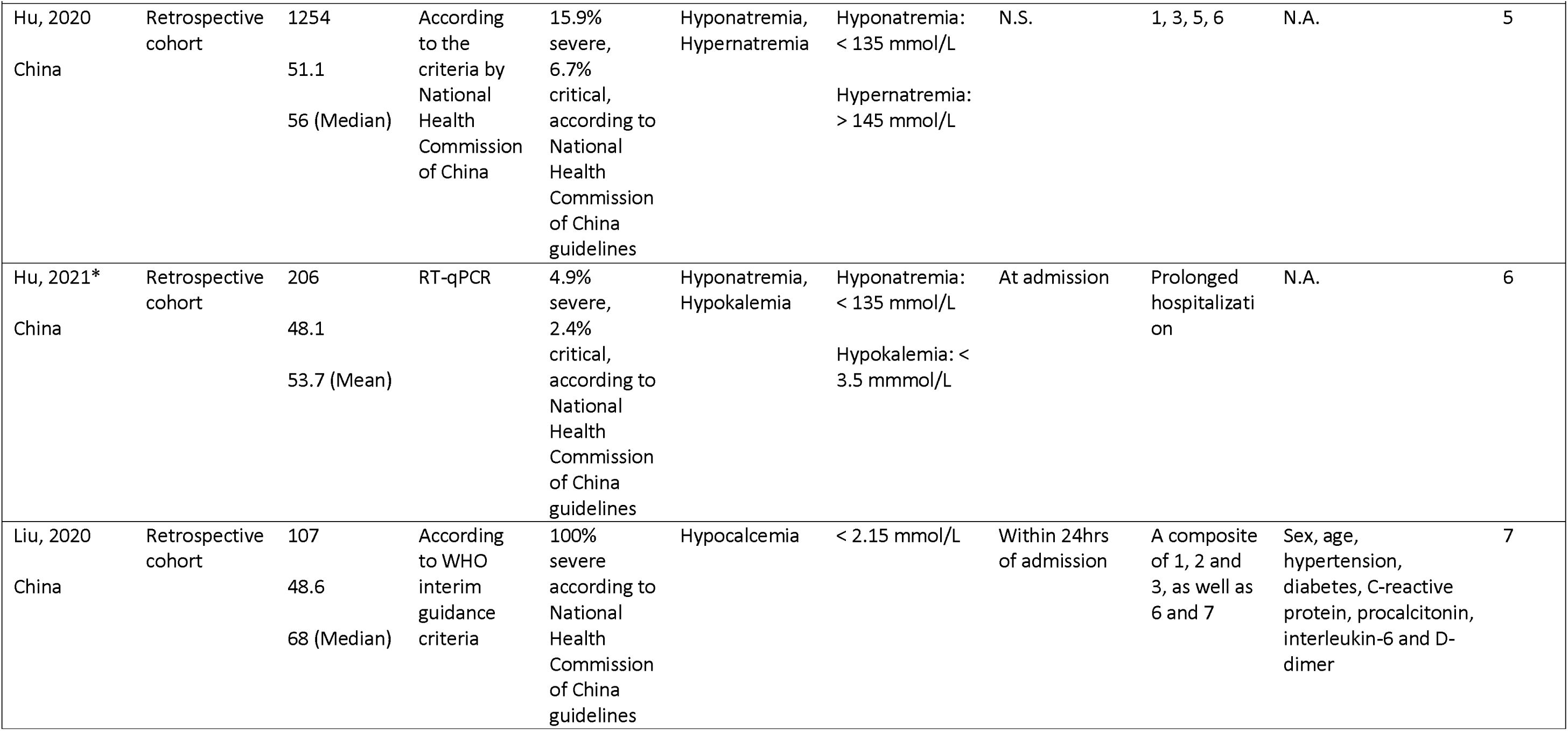
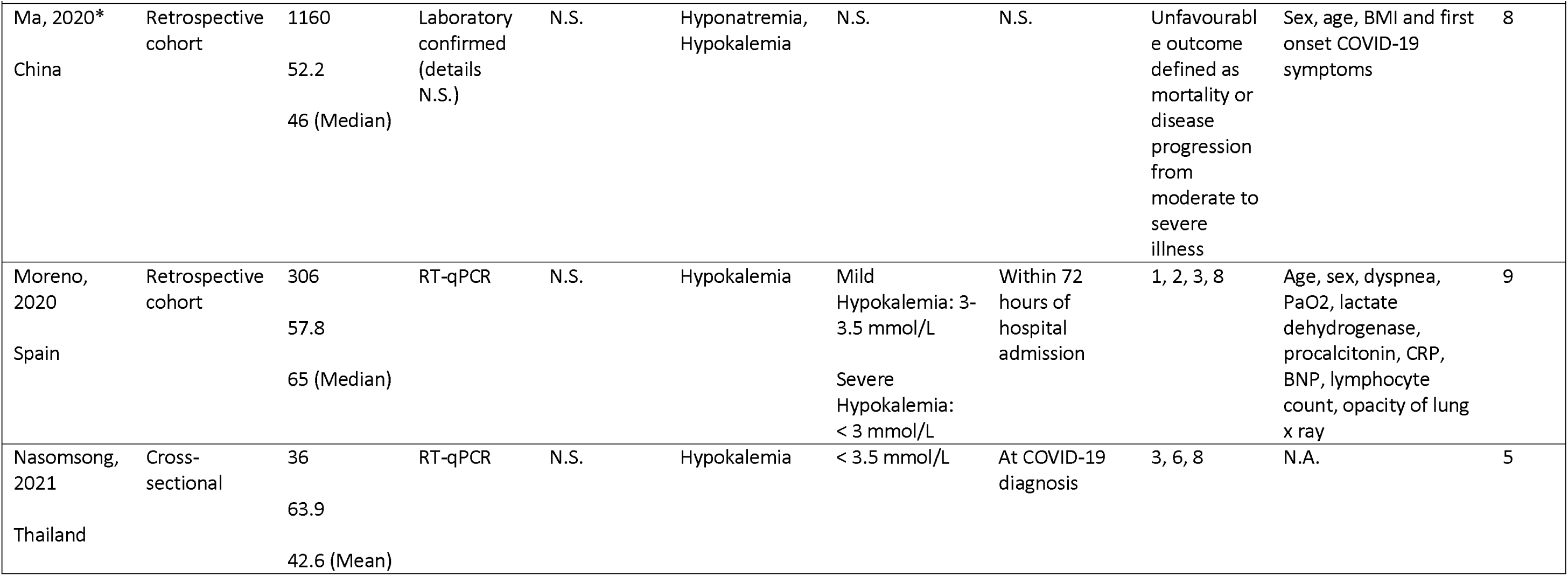
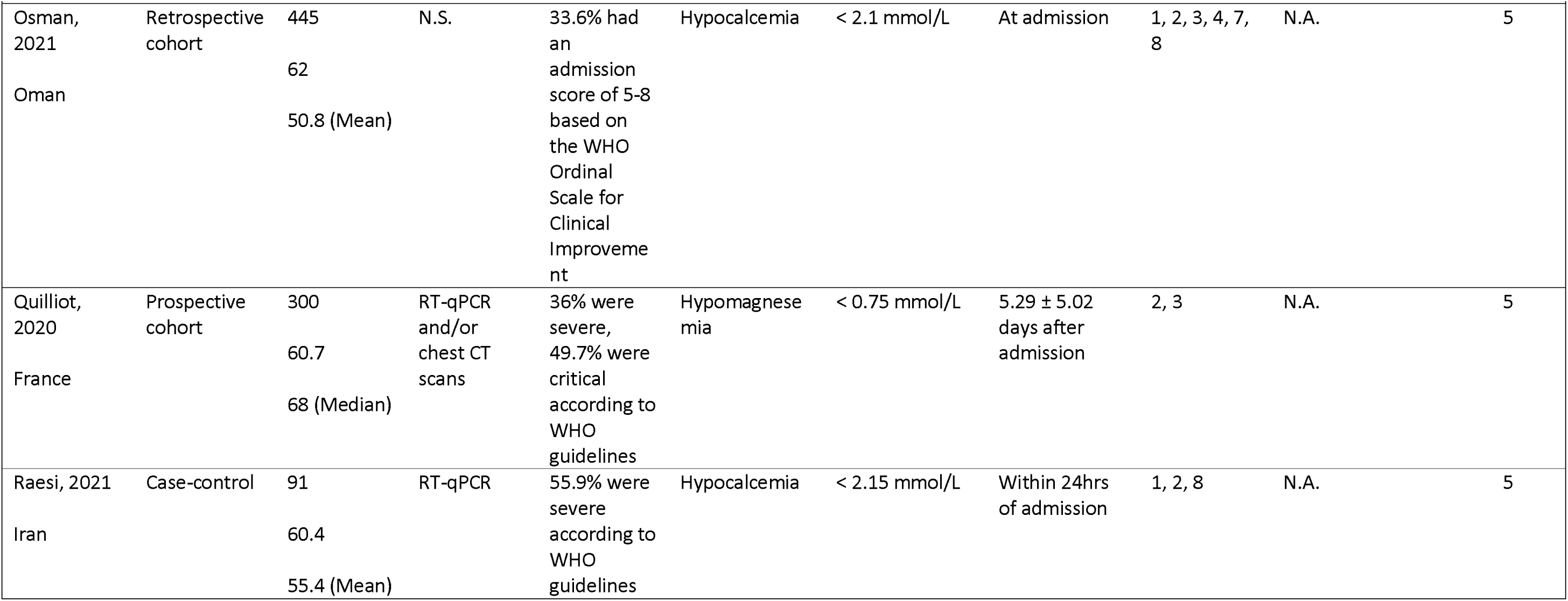
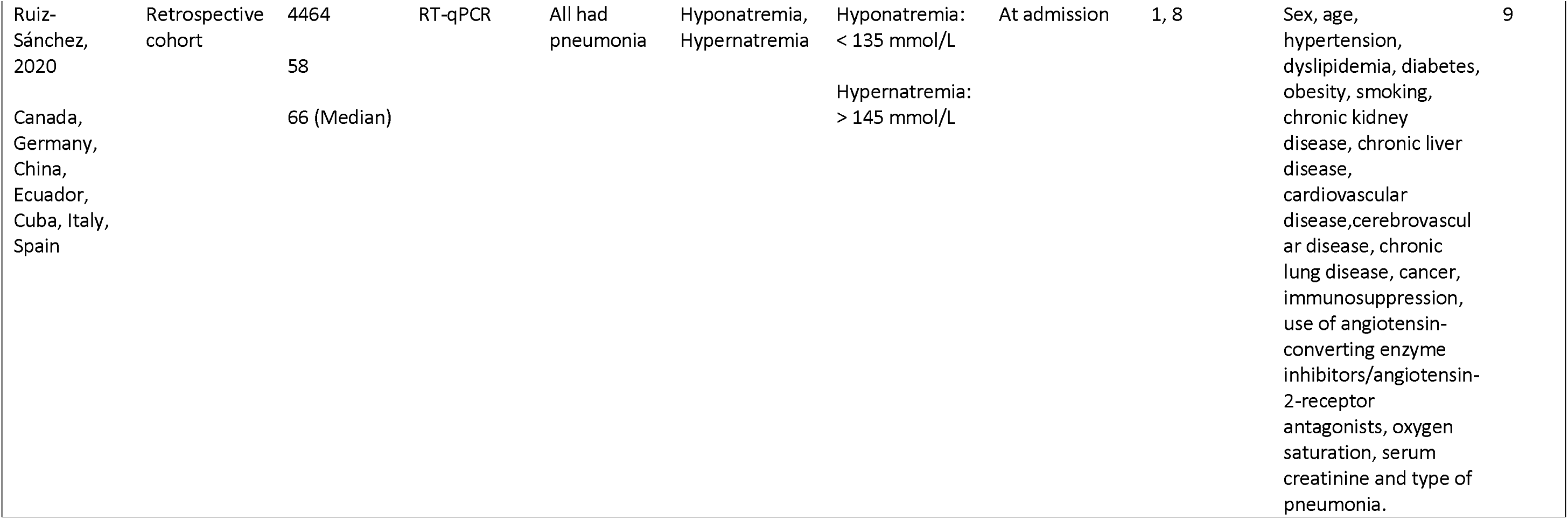
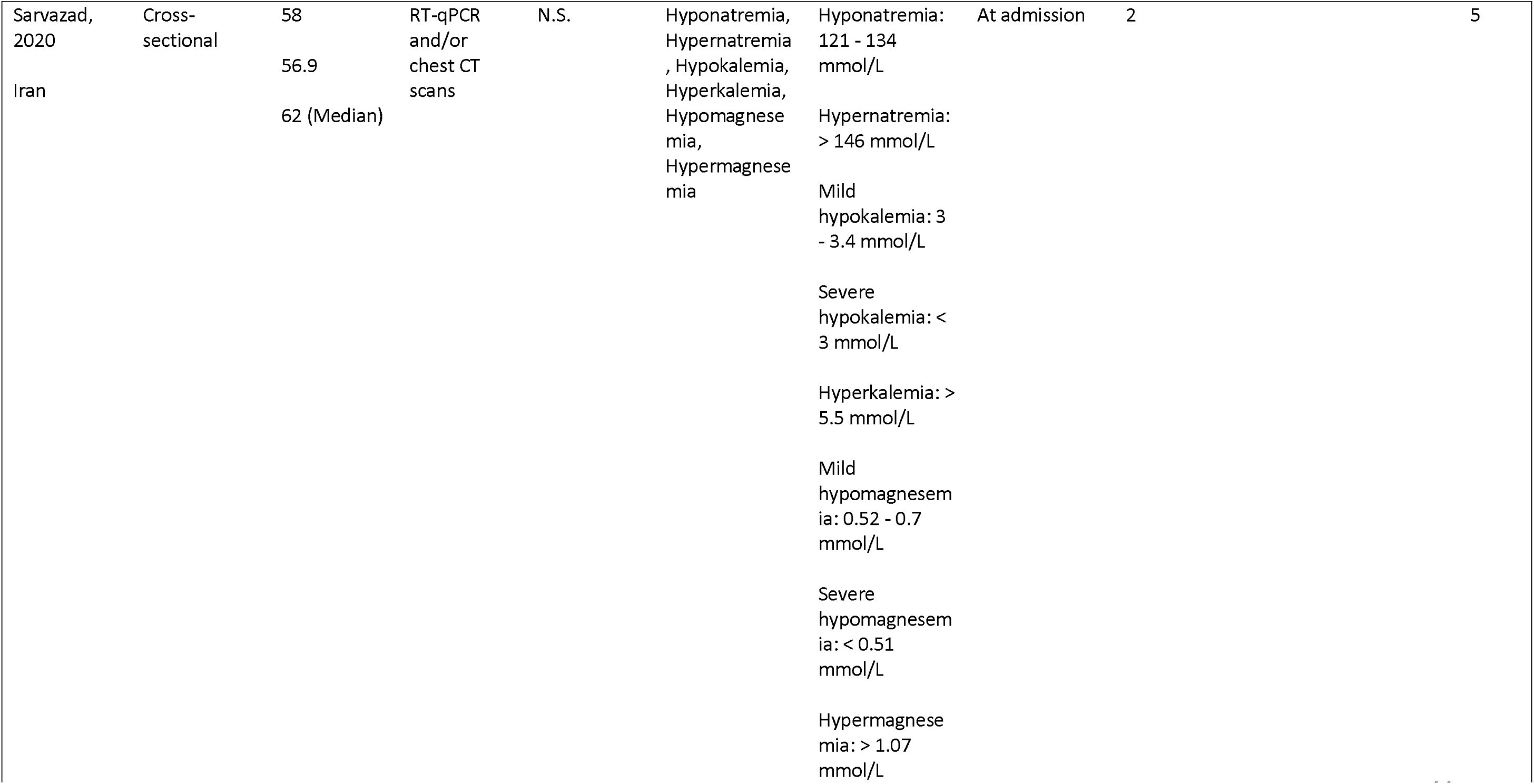
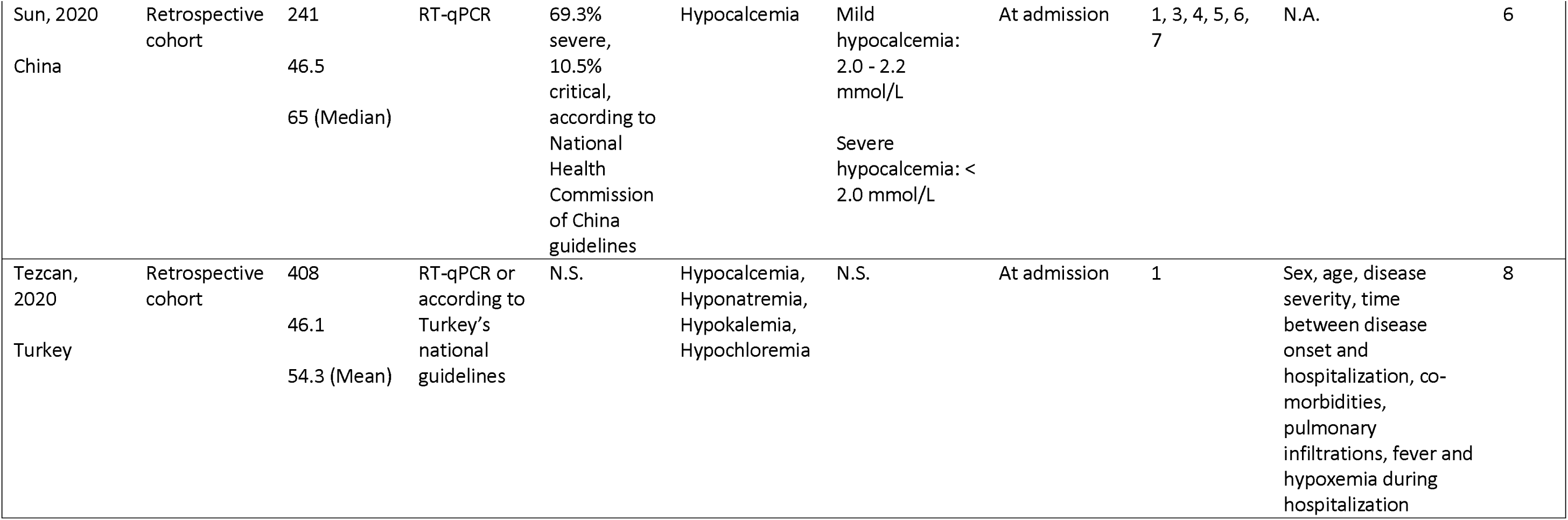
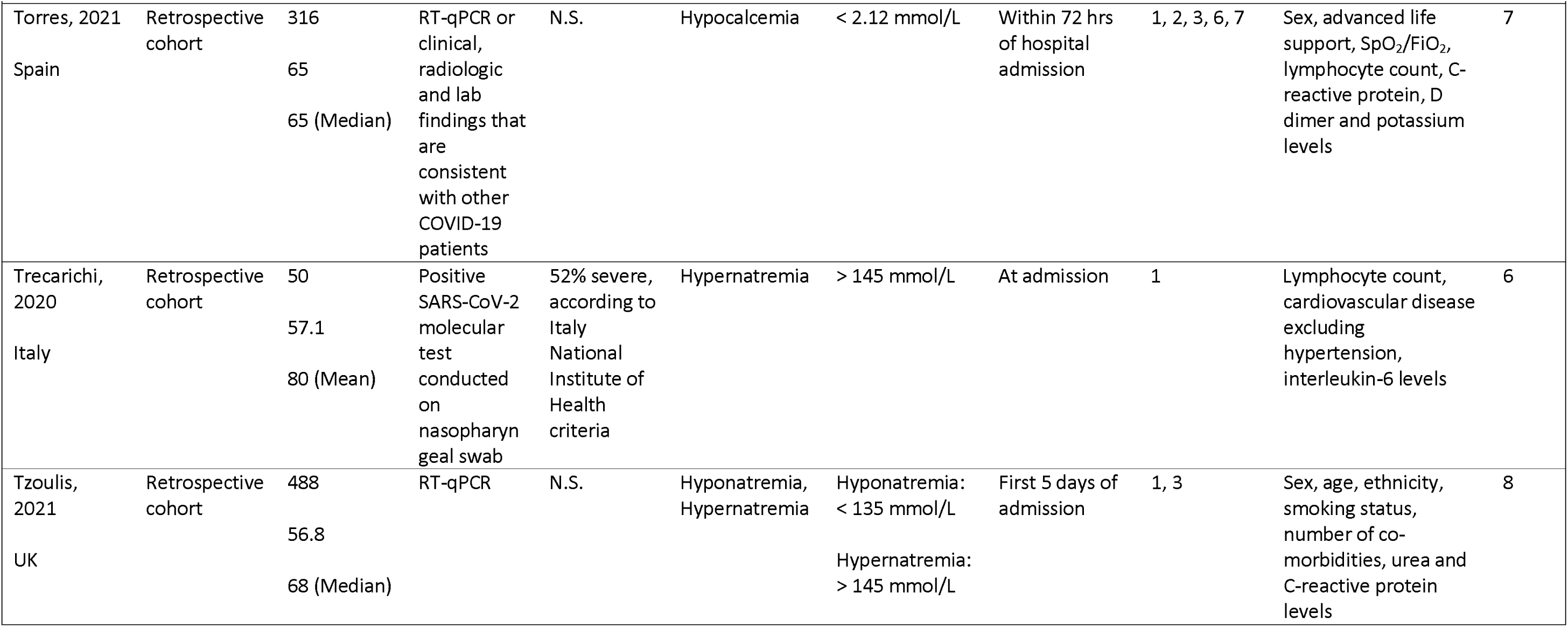
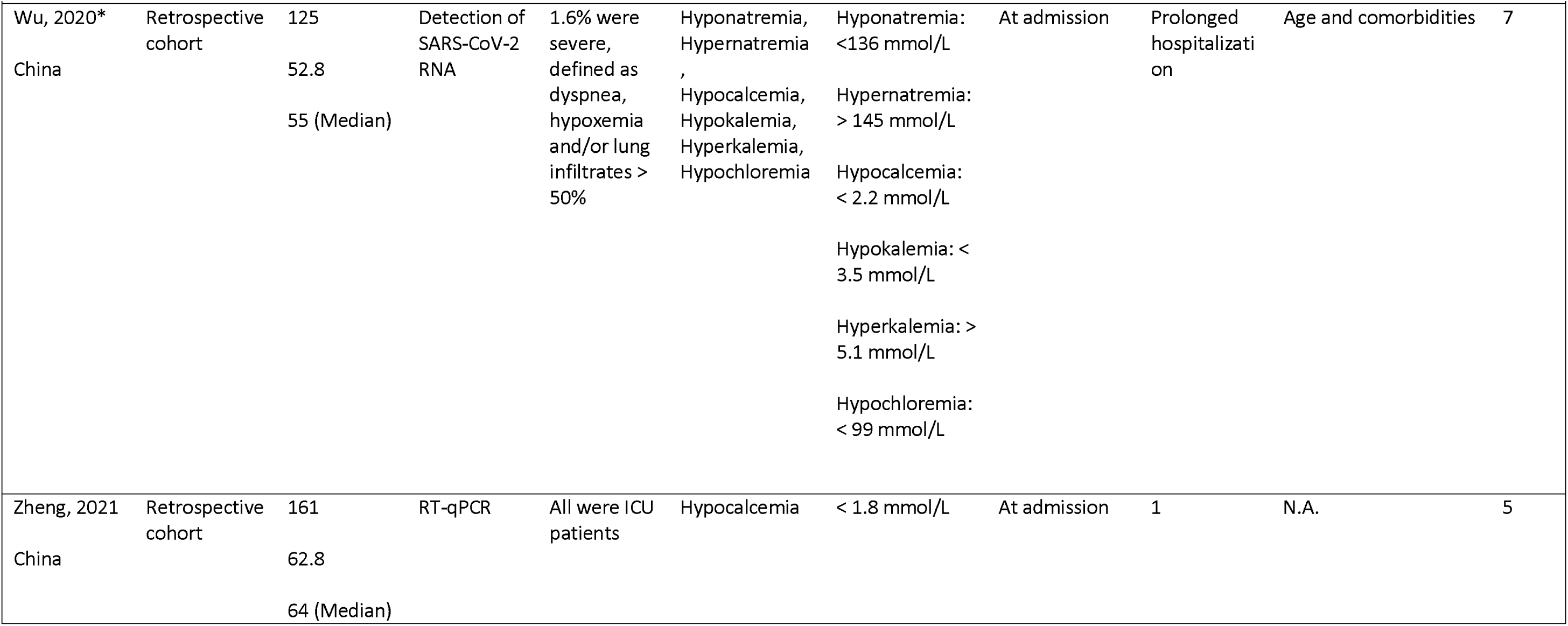
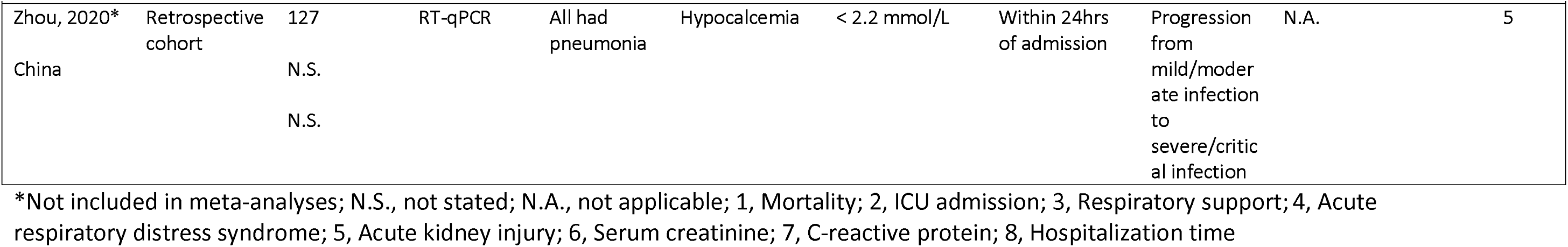
Summary of included studies.

### Definitions of electrolyte imbalances

Studies measured the respective electrolyte levels at hospital admission (15 studies), within 24 hours (4 studies), 72 hours (2 studies) or beyond 72 hours of admission (2 studies). One study measured electrolyte levels at COVID-19 diagnosis and one study recorded the imbalance at any time during hospitalization.[20, 48] As some studies measured electrolyte levels beyond 24 hours,[14, 22, 47, 54] we excluded them in sensitivity analyses. This did not did not alter our conclusions. We searched for but found no studies investigating dysphosphatemia in relation to COVID-19 outcomes.

#### Dysnatremia

Thirteen and ten studies investigated the association of hyponatremia and hypernatremia with the above specified COVID-19 clinical outcomes respectively (**Table 1**).[14, 21, 23, 24, 36, 37, 39, 41–44, 46, 51, 52, 55] Majority of studies defined hyponatremia and hypernatremia as having a serum sodium level of < 135 mmol/L or > 145 mmol/L respectively. Two studies further stratified their sample based on dysnatremia severity.[21, 42] Three studies corrected their sodium measurements with glucose levels.[14, 21, 39]

#### Dyskalemia

Nine and two studies investigated the association of hypokalemia and hyperkalemia with the same COVID-19 outcomes respectively (**Table 1**).[20, 23, 24, 40, 44, 46–48, 52] Majority of studies defined hypokalemia and hyperkalemia as having a serum potassium level of < 3.5 mmol/L or > 5.1 mmol/L respectively. Three studies further stratified their sample based on hypokalemia severity.[40, 47, 52]

#### Dyscalcemia

Ten studies investigated the association of hypocalcemia with the same COVID-19 clinical outcomes (**Table 1**).[23, 24, 38, 45, 49, 50, 53, 54, 56, 57] Majority of the studies defined hypocalcemia as having a serum calcium level of < 2.20 mmol/L, with the exception of one study that defined it as < 1.8 mmol/L.[56] There were no studies that investigated hypercalcemia.

#### Dysmagnesemia

Two studies investigated the association of dysmagnesemia including hypomagnesemia (2 studies) and hypermagnesemia (1 study) with ICU admission and respiratory support (**Table 1**). Hypomagnesemia and hypermagnesemia was defined as having a serum magnesium level of < 0.75 mmol/L or > 1.07 mmol/L respectively.[22, 52]

#### Dyschloremia

Two studies investigated the association of hypochloremia, defined as a serum chloride level of < 99 mmol/L, with prolonged hospitalization and mortality respectively.[23, 24]

### Association of electrolyte imbalances with COVID-19 poor outcome

#### Overall poor outcome

Compared to the control group, participants with hyponatremia (OR=2.08, 95%CI=1.48-2.94, I^2^=93%, N=8), hypernatremia (OR=4.32, 95%CI=3.17-5.88, I^2^=45%, N=7) or hypocalcemia (OR=3.31, 95%CI=2.24-4.88, I^2^=25%, N=6) had, on average, significantly higher pooled odds of poor outcome, defined as a composite of mortality, ICU admission, respiratory support and ARDS (**Figure 2a**). After adjustment, the associations were attenuated but remained significant for hyponatremia (aOR=1.65, 95%CI=1.09-2.51, I^2^=91%, N=5) and hypernatremia (aOR=2.10, 95%CI=1.80-2.44, I^2^=0%, N=3) (**Figure 2b**). There was no significant association found for participants with hypokalemia (OR=0.96, 95%CI=0.62-1.51, I^2^=0%, N=4) or hypomagnesemia (OR=1.43, 95%CI=0.21-9.60, I^2^=86%, N=2) (**Figure 2a**). Between-study heterogeneity was significant for hyponatremia (I^2^=91%) and hypomagnesemia (I^2^=86%) but expected due to the pooling of different clinical outcomes. As ICU admission criteria differ across countries, we performed sensitivity analyses excluding ICU admission, which did not change our findings. In studies excluded from meta-analysis, Ma et al. reported participants with hyponatremia and/or hypokalemia having an increased odds of unfavourable outcome, defined as mortality or disease progression from moderate to severe (OR=19.44, 95%CI=11.47-32.96), after adjusting for sex, age, BMI and first-onset COVID-19 symptoms.[46] Zhou et al. reported participants with low calcium levels tended to progress to a severe or critical infection.[57]

**Figure 2:**
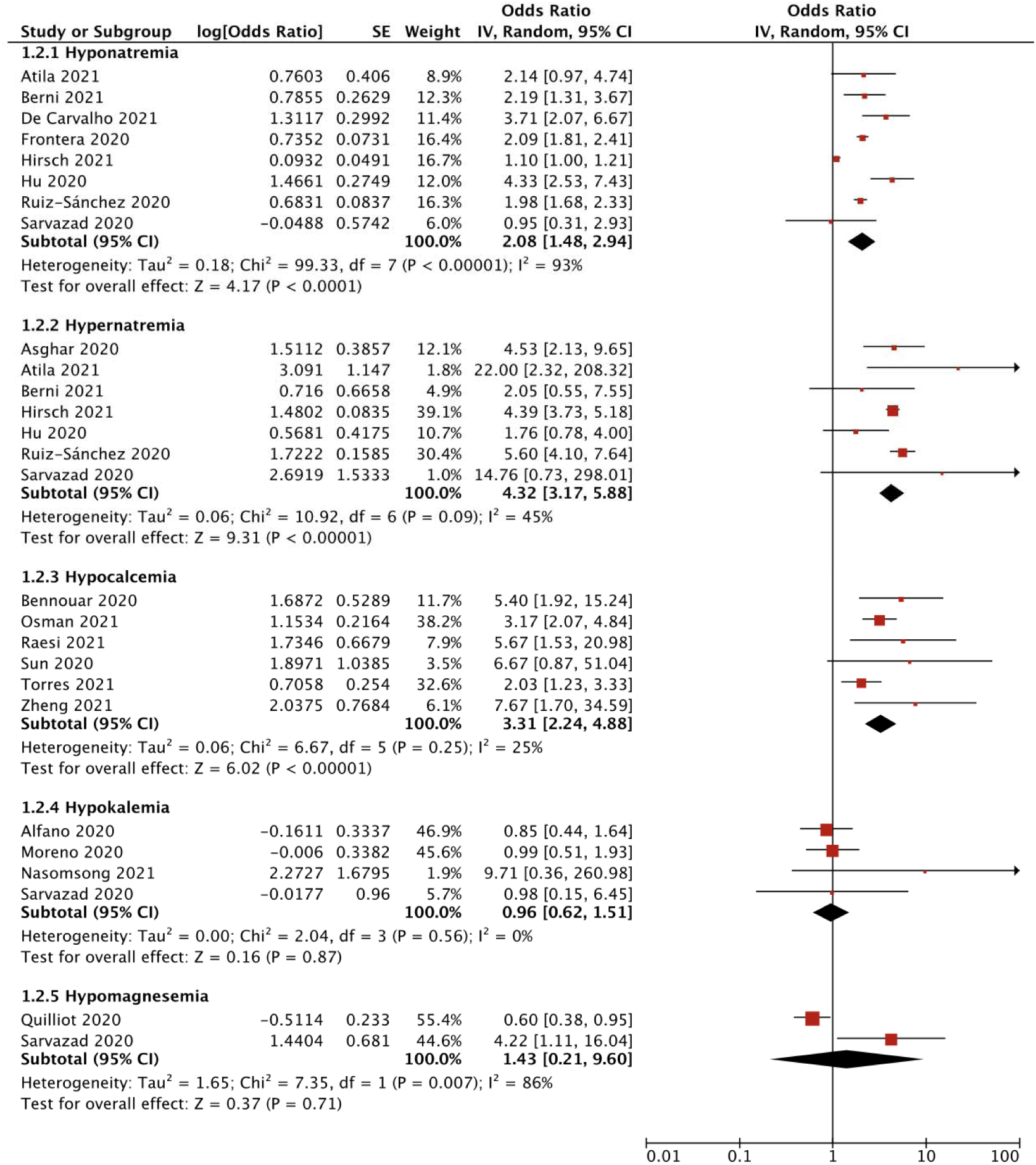
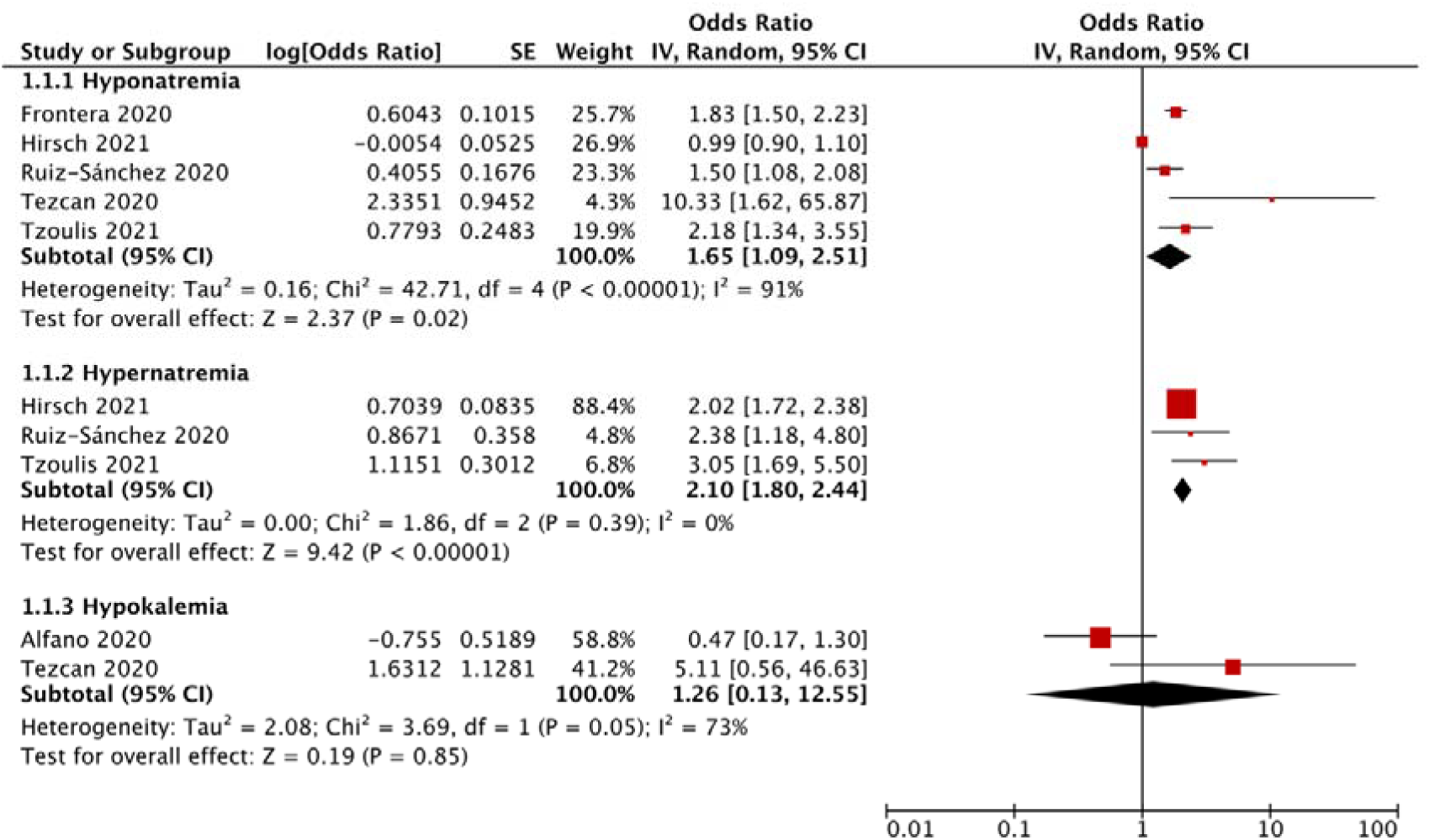
Forest plot showing the (a) unadjusted and (b) adjusted association between electrolyte imbalances with poor outcome*, stratified by the type of electrolyte imbalance. Legend: Black diamonds are the estimated pooled odds ratios for each random-effects meta-analysis; red boxes reflect the relative weight apportioned to studies in the meta-analysis *Poor outcome Is defined as a composite of mortality, ICU admission, respiratory support and acute respiratory distress syndrome Figure 2a: Poor outcome, unadjusted association, stratified by the type of electrolyte imbalance Figure 2b: Poor outcome, adjusted association, stratified by the type of electrolyte imbalance

#### Mortality

Looking at the specific poor outcome composites, participants with hyponatremia (OR=2.15, 95%CI=1.46-3.17, I^2^=94%, N=7), hypernatremia (OR=5.60, 95%CI=3.57-8.78, I^2^=73%, N=6) or hypocalcemia (OR=2.72, 95%CI=1.34-5.51, I^2^=64%, N=6) had, on average, significantly higher pooled odds of mortality compared to the control group (**Supplemental Figure S1a**). The adjusted association remained significant for hyponatremia (aOR=1.48, 95%CI=1.03-2.12, I^2^=75%. N=7) and hypernatremia (aOR=3.32, 95%CI=1.79-6.15, I^2^=82%, N=5) (**Supplemental Figure S1b**). There were insufficient studies that calculated the adjusted association for hypocalcemia. There were no significant associations found for participants with hypokalemia (OR=0.92, 95%CI=0.57-1.46, I^2^=0%). Pre-specified sensitivity analyses excluding studies that did not measure electrolyte imbalance at admission,[43, 54] moderate risk-of-bias studies[41, 43] and non-cohort studies did not change our results substantially.[50] However, pre-specified sensitivity analyses excluding studies that corrected sodium levels with glucose concentration decreased between-study heterogeneity for hyponatremia (I^2^=37%).[14, 21, 39] In a study excluded from meta-analysis, Tezcan et al. reported no significant association between hypochloremia and mortality (OR=2.60, 95%CI=0.28-23.54).[23]

#### ICU Admission

Compared to the control group, participants with hyponatremia (OR=2.19, 95%CI=1.36-3.52, I^2^=44%, N=4) or hypocalcemia (OR=2.23, 95%CI=1.60-3.11, I^2^=6%, N=3) had on average, significantly higher odds of ICU admission. There were no significant associations for hypernatremia (OR=3.72, 95%CI=0.14-99.22, I^2^=82%, N=3), hypokalemia (OR=1.35, 95%CI=0.25-7.30, I^2^=90%, N=3) or hypomagnesemia (OR=1.43, 95%CI=0.21-9.61, I^2^=86%, N=2) (**Supplemental Figure S3**). Insufficient studies calculated the adjusted association for meta-analysis.

#### Respiratory Support

A total of 12 studies reported the use of respiratory support, defined as the need for either invasive ventilation or non-invasive ventilation.[14, 36, 37, 39, 41–43, 47–49, 53, 54] Compared to the control group, participants with hyponatremia (OR=2.16, 95%CI=1.91-2.45, I^2^=0%, N=5), hypernatremia (OR=3.24, 95%CI=1.24-8.50, I^2^=70%, N=4), hypocalcemia (OR=2.99, 95%CI=2.16-4.14, I^2^=0%, N=3) or hypokalemia (OR=5.69, 95%CI=2.81-11.54, I^2^=0%, N=2) had on average, a significantly higher odds of requiring respiratory support. The adjusted association remained significant for hyponatremia (aOR=1.88, 95%CI=1.56-2.26, I^2^=0%, N=2) (**Supplemental Figure S4b**). There were insufficient studies that calculated the adjusted association for hypernatremia, hypercalcemia and hypokalemia. Between-study heterogeneity for participants with hypernatremia was eliminated (I^2^=0%) and the association was attenuated after exclusion of studies that adjusted sodium levels for glucose concentration during pre-specified sensitivity analyses (OR=4.61, 95%CI=3.01-7.08).[39] Further subgroup analyses by the type of ventilation for hypernatremic studies eliminated between-study heterogeneity in the invasive ventilation subgroup (OR=6.73, 95%CI=1.40-32.37, I^2^=0%, N=2) and the association was no longer significant in the non-invasive ventilation subgroup (OR=0.77, 95%CI=0.05-13.12, I^2^=83%, N=2) (**Supplemental Figure S5b**). The association remained significant with low between-study heterogeneity in both invasive ventilation and non-invasive ventilation for hyponatremic studies (**Supplemental Figure S5a**)

#### ARDS

Compared to the control group, participants with hypocalcemia (OR=2.41, 95%CI=0.96-6.08, I^2^=27%, N=2) (**Supplemental Figure S6**) had an increased odds of ARDS, although this was not statistically significant. In studies excluded from the meta-analysis, Atila et al. reported participants with hyponatremia (OR=2.35, 95%CI=0.89-6.19) and hypernatremia (OR=16.05, 95%CI=2.39-107.63) had an increased odds of developing ARDS.[37]

### Performance and prognostic value of electrolyte imbalances in predicting unadjusted odds of poor outcome

Based on the SROC curves generated for each electrolyte imbalance, hypernatremia (AUC=0.80, 95%CI=0.76-0.83) and hypocalcemia (AUC=0.71, 95%CI=0.67-0.75) performed adequately, with AUC >0.70. In particular, hypernatremia was 97% specific (95%CI=0.94-0.98; LR+ 4.0) for a poor outcome with low sensitivity (0.13, 95%CI=0.07-0.22; LR- 0.90), while hypocalcemia was 76% sensitive (95%CI=0.53-0.90; LR- 0.44) for a poor outcome with low specificity (0.53, 95%CI=0.26-0.78; LR+ 2.0) (**Figure 3a and Figure 4a**). In contrast, hyponatremia and hypokalemia performed inadequately (AUC <0.70) (**Supplemental Results**). Visual inspection of coupled funnel plots did not indicate a clear threshold effect (**Figure 3c** and **Figure 4c**).

**Figure 3:**
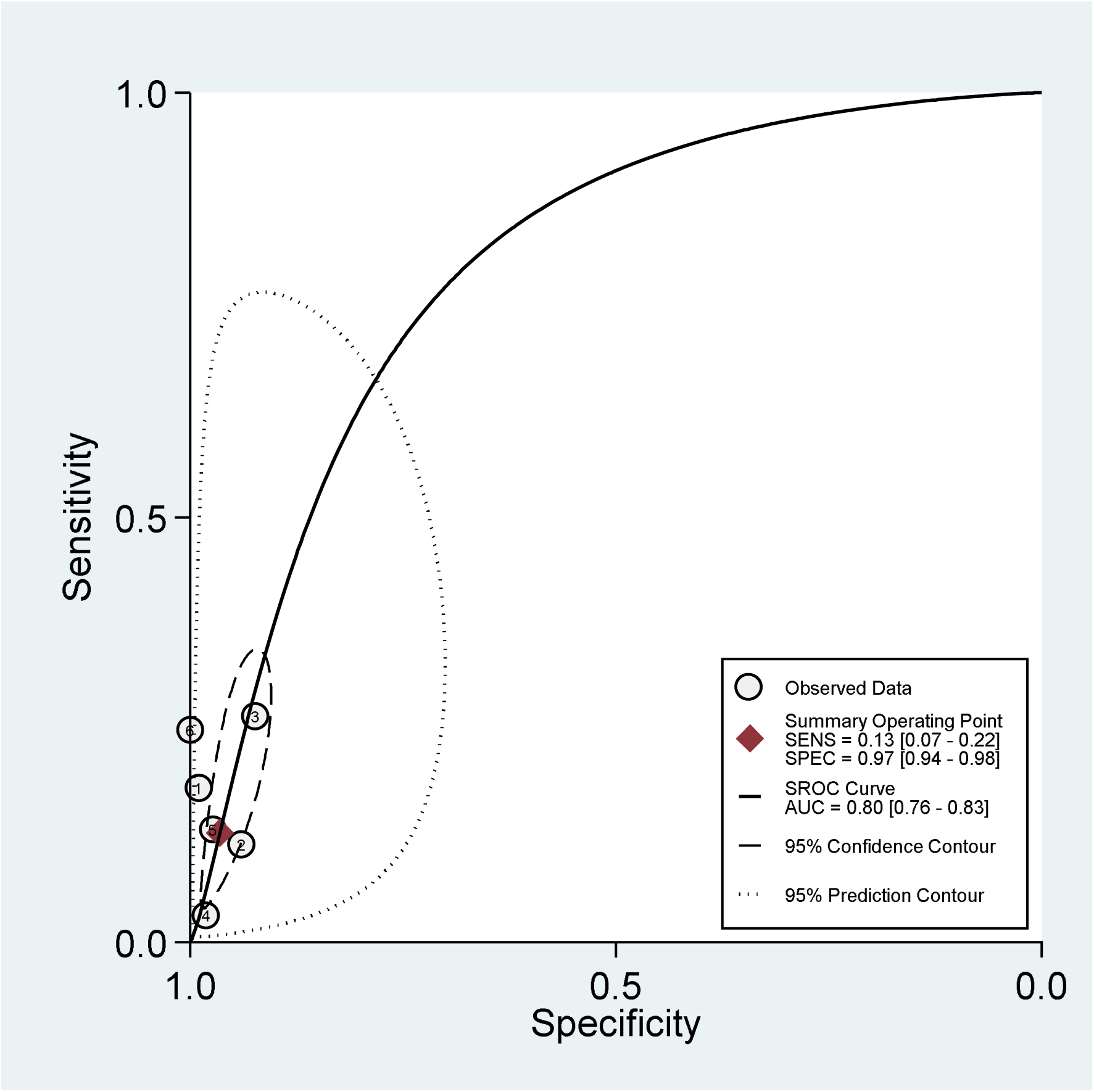
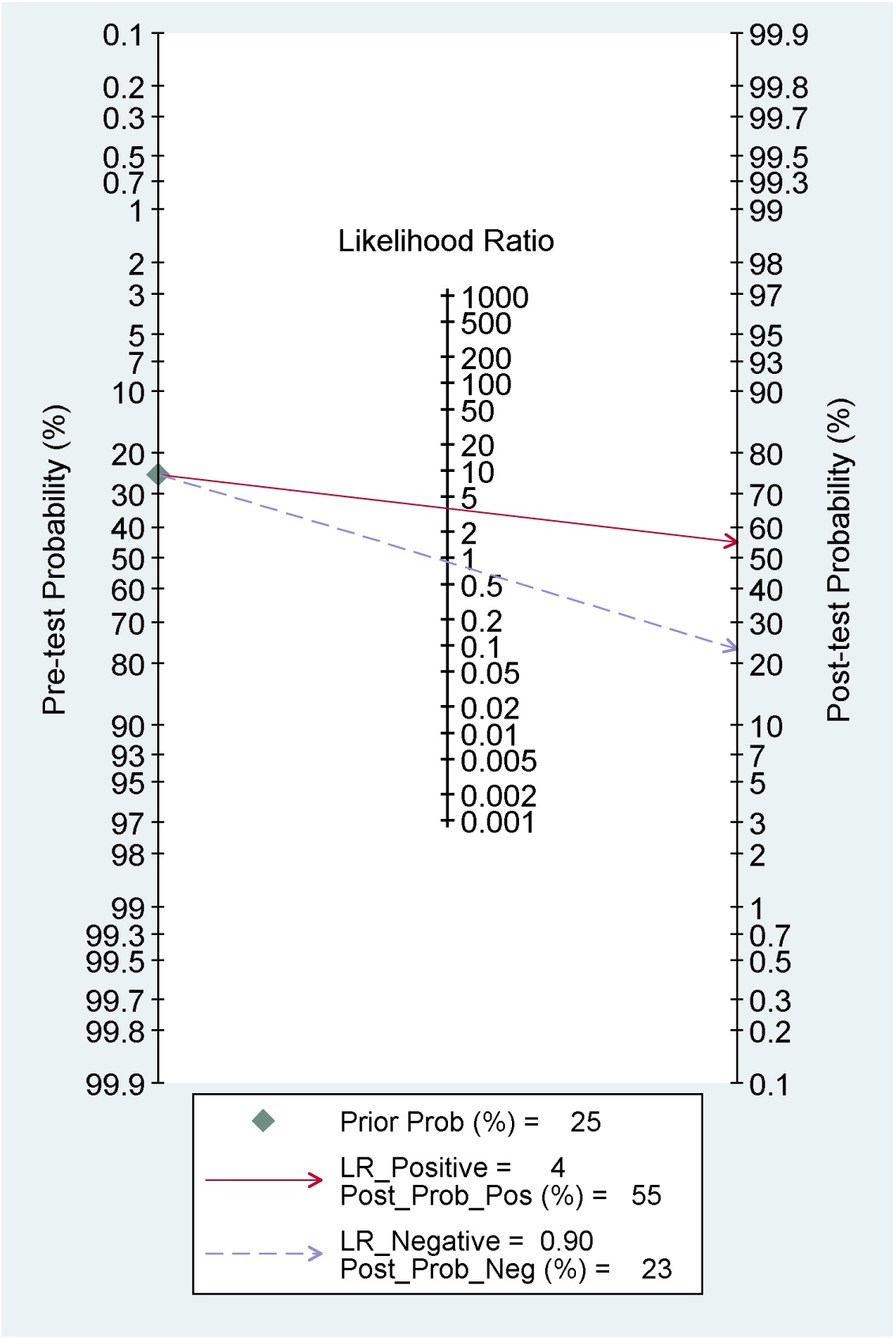
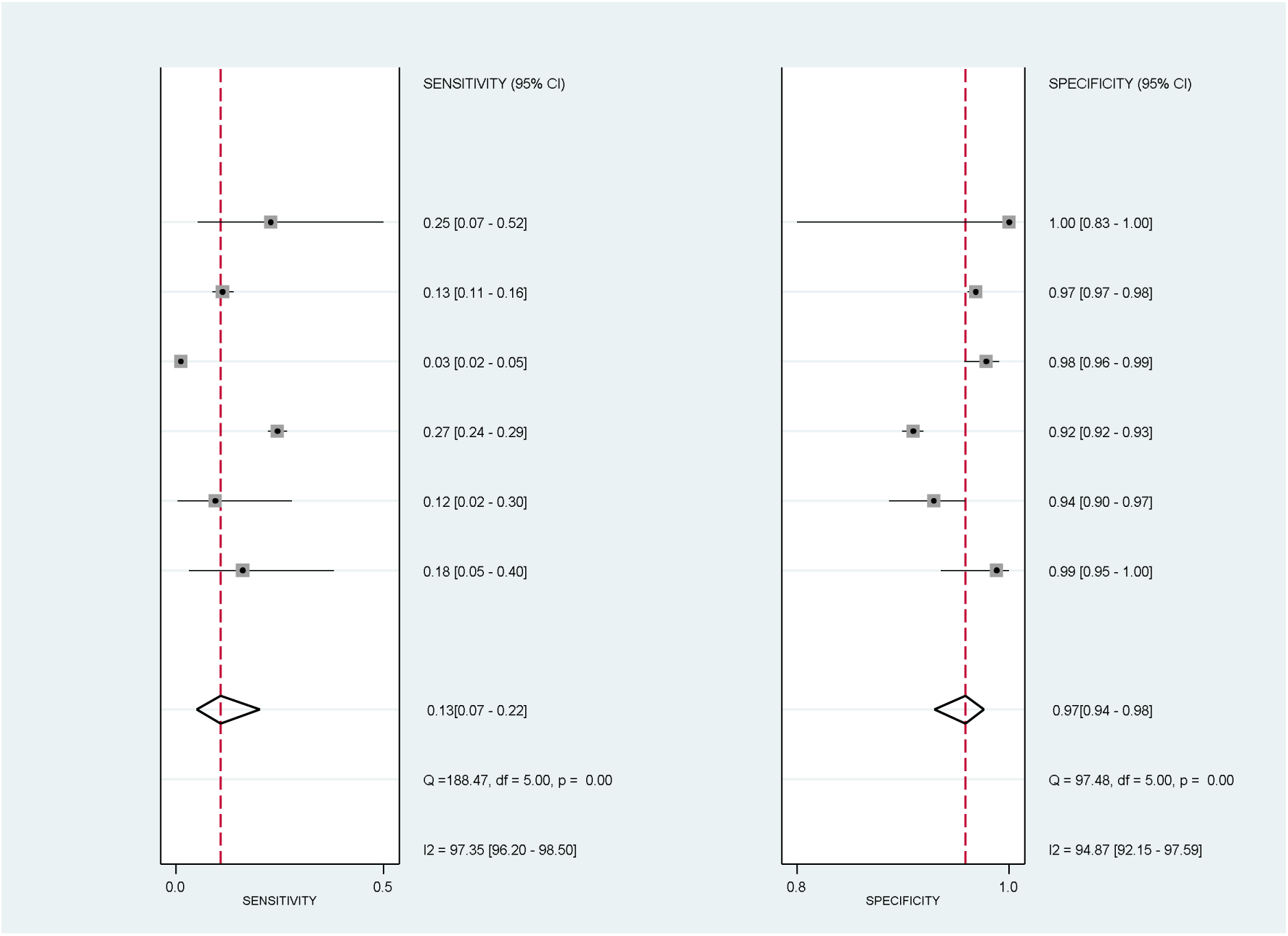
(a) Summary Receiver Operator Characteristic Curve, (b) Fagan plot and (c) Coupled funnel plot of hypernatremia in predicting poor outcome. Figure 3a: Summary Receiver Operator Characteristic Curve of hypernatremia and poor outcome Figure 3b: Fagan plot of hypernatremia and poor outcome Figure 3c: Coupled funnel plot of hypernatremia and poor outcome

**Figure 4:**
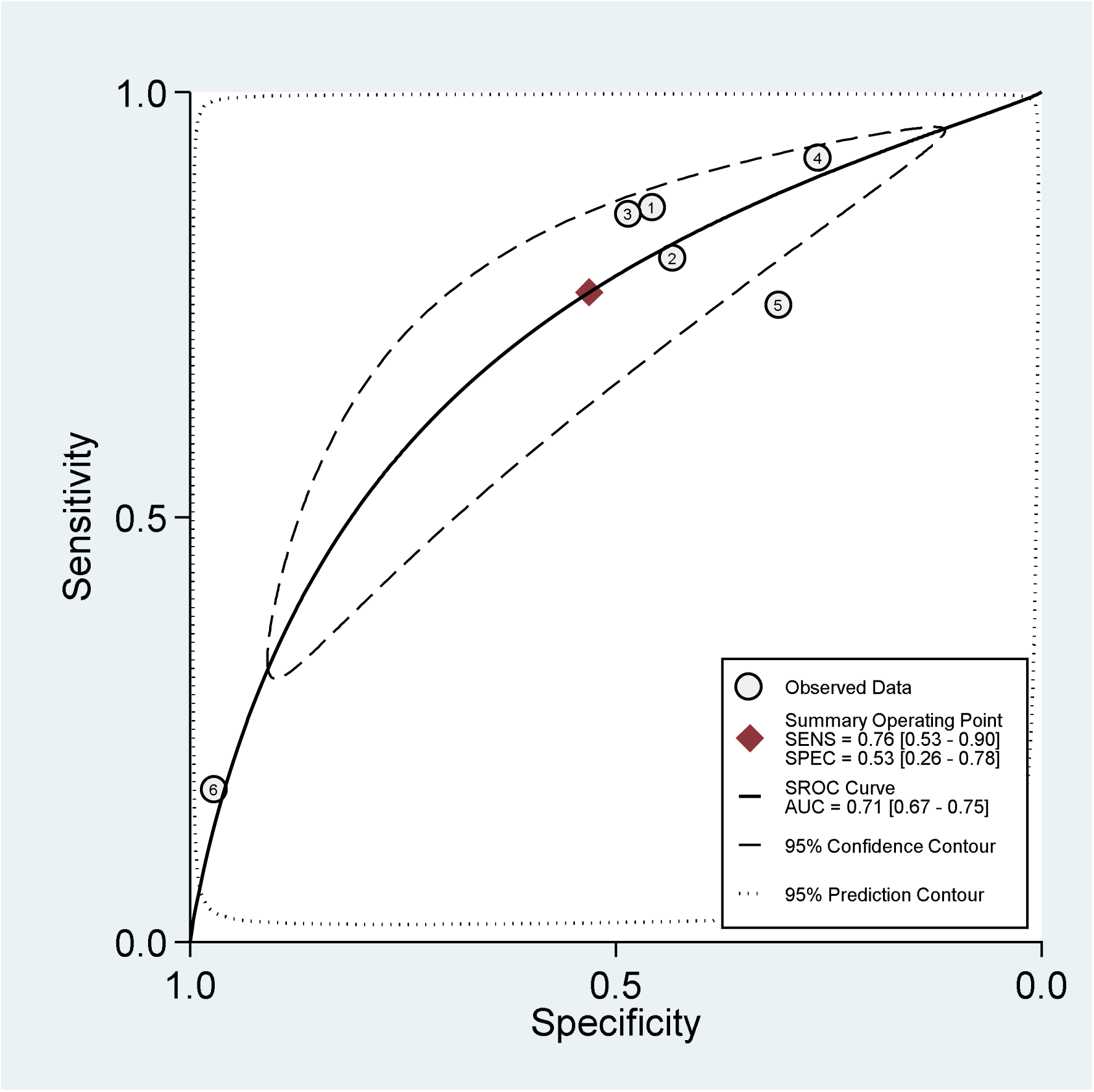
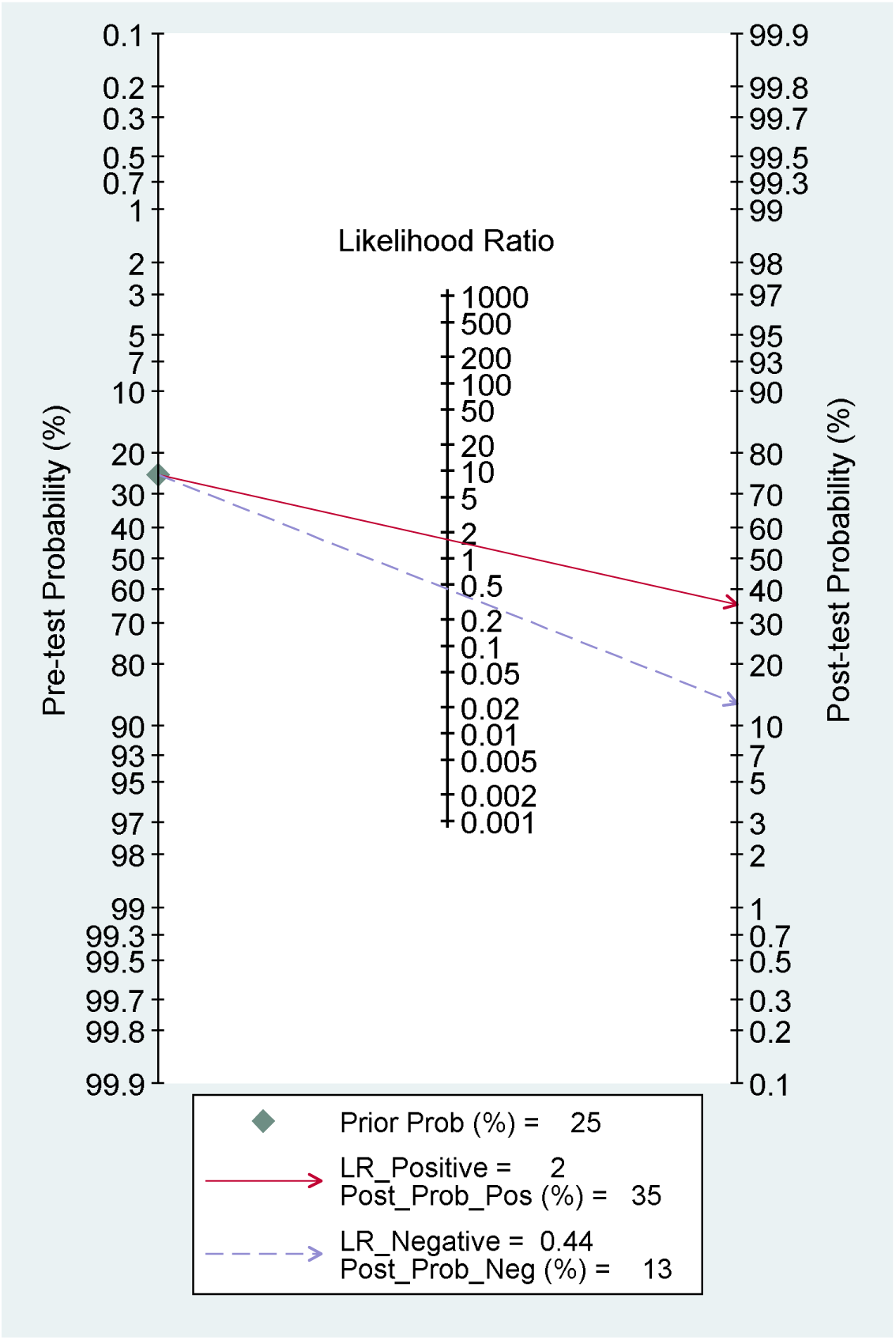
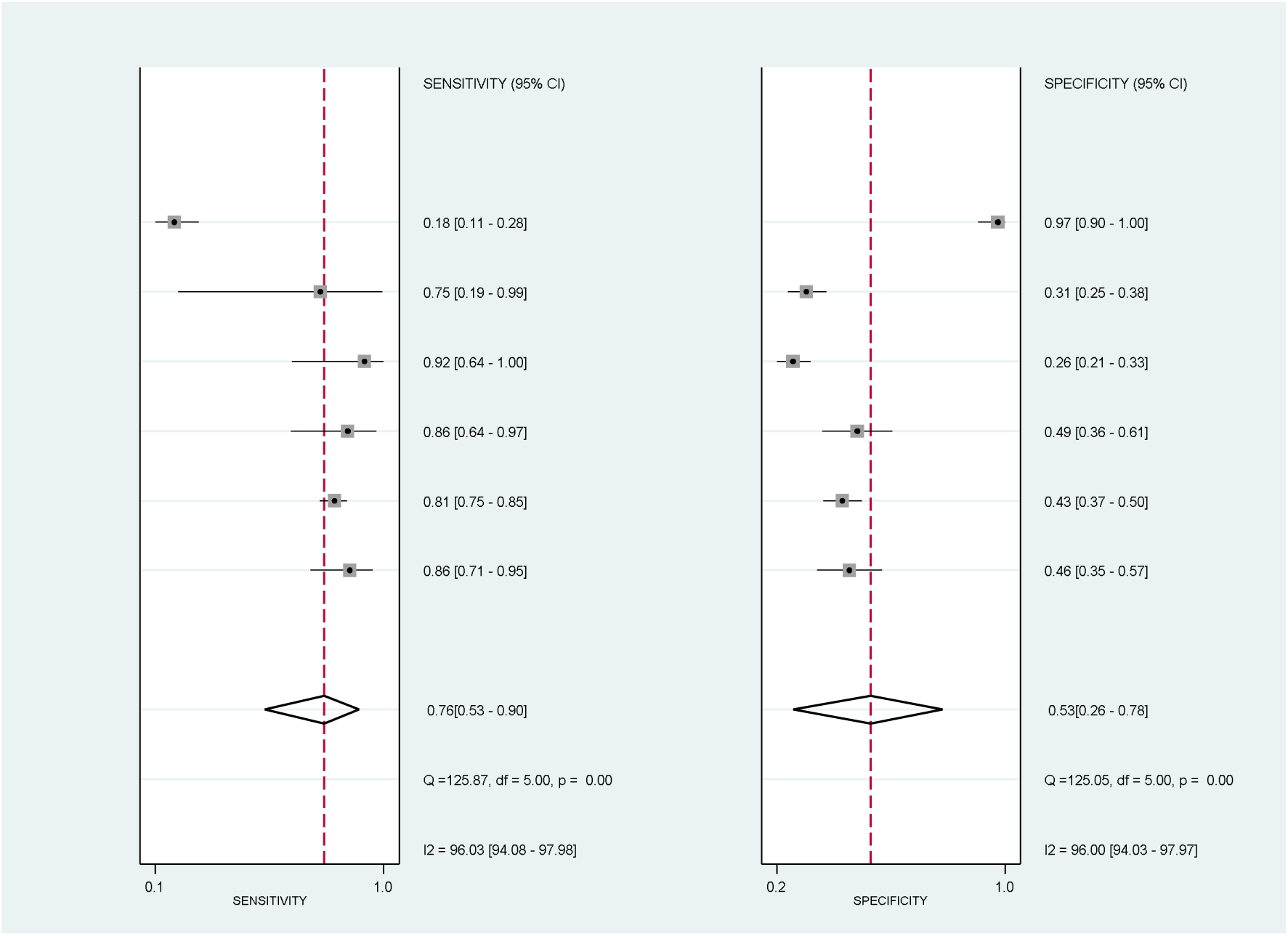
(a) Summary Receiver Operator Characteristic Curve, (b) Fagan plot and (c) Coupled funnel plot of hypocalcemia in predicting poor outcome. Figure 4a: Summary Receiver Operator Characteristic Curve of hypocalcemia and poor outcome Figure 4b: Fagan plot of hypocalcemia and poor outcome Figure 4c: Coupled funnel plot of hypernatremia and poor outcome

Assuming a 25% pre-test probability of progression to severe COVID based on published estimates in the general population infected with COVID,[58–60] the presence of hypernatremia (LR+ 4.0) would be associated with a PPV of 55%, or a 55% post-test probability of progression to severe COVID based on Fagan’s nomogram (**Figure 3b**). Similarly, the absence of hypocalcemia (LR- 0.44) would be associated with a 13% post-test probability of severe COVID, or a NPV of 87% (**Figure 4b**).

### Association of electrolyte imbalances with Acute Kidney Injury (AKI) and serum creatinine levels

Participants with hyponatremia had a significantly increased odds ratio (OR=1.63, 95%CI=1.26-2.10, I^2^=13%, N=2) (**Supplemental Figure S7**) compared to the controls. In studies excluded from the meta-analysis, Sun et al. and Alfano et al. reported no significant association for hypocalcemia (OR=4.67, 95%CI=0.59-36.47) and hypokalemia (OR=0.88, 95%CI=0.49-1.60) respectively.[14, 20, 53] Additionally, Tzoulis et al. concluded that sodium values were not associated with the risk for AKI, although sufficient data was not provided. Compared to the control group, there was no significant difference in serum creatinine levels for dysnatremia, hypocalcemia and hypokalemia (**Supplemental Figure S8a**).

### Association of electrolyte imbalances with C-reactive protein (CRP) levels

While hyponatremia (MD=27.92mg/L, 95% CI = 16.97 to 38.86mg/L, I^2^=56%, N=3), hypocalcemia (MD=10.18mg/L, 95% CI = 7.15 to 13.20mg/L, I^2^=0%, N=4) and hypokalemia (MD=5.82mg/L, 95% CI = 0.26 to 11.37mg/L, I^2^=0%, N=2) showed significantly higher CRP levels as compared to the control group, hypernatremia (MD=57.16mg/L, 95% CI = -27.12 to 141.45mg/L, I^2^=98%, N=3) showed no significant difference (**Supplemental Figure S8b**).

### Association of electrolyte imbalances with Interleukin-6 (IL-6) levels

In studies excluded from meta-analyses, Berni et al. reported significantly higher (p-value <0.001) baseline IL-6 levels in hyponatremic participants as compared to the control group. There was no significant difference in IL-6 levels between hypernatremic and normonatremic participants (p-value=0.395).[39] Liu et al. reported a significantly higher IL-6 levels (p-value=0.0276) in hypocalcemic participants as compared to the control group.[45]

### Association of electrolyte imbalances with hospitalization time

Compared to the control group, participants with hypocalcemia (MD=2.25, 95% CI = 0.10 to 4.40, p=0.04, I^2^=70%, N=2) had a significantly longer hospitalization time while participants with hyponatremia (MD=2.45, 95% CI = -0.98 to 5.88, p=0.16, I^2^=93%, N=4), hypernatremia (MD=1.59, 95% CI = -5.57 to 8.74, p=0.66, I^2^=82%, N=3) and hypokalemia (MD=0.87, 95% CI = -8.34 to 10.08, p=0.85, I^2^=93%, N=2) showed no significant difference. In studies excluded from meta-analyses, Wu et al. reported participants with hypochloremia (OR=2.67, 95%CI=1.10-6.62) and hypocalcemia (OR=3.31, 95%CI=1.39-7.89) as having a significantly higher odds of long-term hospitalization, defined as ≥ 14 days, as compared to the control group.[24] Tzoulis et al. reported no significant associations with prolonged hospitalization, defined as ≥ 7 days, for hyponatremia (OR=1.09, 95%CI=0.96-1.21) and hypernatremia (OR=1.37, 95%CI=0.70-2.65).[14]

## DISCUSSION

In this systematic review and meta-analysis of 28 observational studies comprising a combined cohort of 26,897 participants with COVID-19, we found that hyponatremia, hypernatremia and hypocalcemia were associated with a 2-fold, 4-fold and 3-fold increased odds of poor clinical outcome, defined as a composite of mortality, ICU admission, ARDS and respiratory support. Participants with hyponatremia had a 63% increased odds of AKI. Hypernatremia and hypocalcemia performed adequately with an AUC score of more than 0.7. Hypernatremia had a specificity of 97% and hypocalcemia had a sensitivity of 76%, suggesting their predictive utility for a poor clinical outcome. The association of hypernatremia and poor outcome could not be explained by differences in CRP and IL-6 compared to normonatremic controls, thus highlighting its potential use as a unique clinical indicator of disease progression. These associations were robust to pre-specified sensitivity analyses and attenuated but remained significant upon adjustment for covariates. Hypokalemia, hypomagnesemia and hypochloremia was not significantly associated with a poor outcome and AKI.

To the best of our knowledge, this is the first comprehensive systematic review and meta-analysis looking at multiple electrolyte imbalances and its associations with a poor clinical outcome. Our findings are consistent with recent meta-analyses that also reported significantly higher odds of poor outcome and severe infection amongst hyponatremic and hypocalcemic participants respectively.[18, 19] We further add value to these studies by including ARDS and AKI as additional outcomes as well as by investigating additional electrolyte imbalances – hypernatremia, hypokalemia, hypochloremia and hypomagnesemia.

These associations could likely be confounded by the underlying disease process – either as part of a non-specific septic response or via mechanisms specific to COVID-19. Marked elevation of inflammatory cytokines have been described in COVID-19, leading in certain instances to cytokine storms.[61] This increase in cytokines can result in syndrome of inappropriate secretion of antidiuretic hormone (SIADH) via inflammatory cytokines, such as IL-6, directly stimulating non-osmotic release of anti-diuretic hormone (ADH) or via injuring lung tissue and alveolar cells inducing SIADH via the hypoxic pulmonary vasocontriction pathway.[62–64] This was observed in our results where hyponatremia, hypocalcemia and hypokalmia were significantly associated with higher baseline CRP levels, which itself is an established marker for inflammation and a more severe disease course. [65] Electrolyte imbalances may also be a general indication of kidney involvement where AKI has been reported to be prevalent in patients hospitalized with COVID-19.[66]

However, in a number of the included primary studies, the association of some electrolyte imbalances with poorer outcome in COVID-19 remained significant even after adjusting for inflammatory biomarkers. Furthermore, our study found that CRP levels were not significantly different between hypernatremic and normonatremic patients, which could suggest that the poor outcome associated with hypernatremia may be unrelated to the systemic inflammatory response. It has been proposed that hypernatremia can be caused by increased angiotensin II activity secondary to SARS-CoV-2-induced down regulation of ACE2 receptors in the proximal tubule after viral entry. [67] While hypernatremia in COVID disease likely represents dehydration from insensible water losses such as fever and tachypnea, it is not clear if dehydration alone can explain the observed poor prognosis, as a recent case series documented persistent hypernatremia despite adequate infusion of free water in 6 patients. [68] This could be consistent with a COVID-19-specific mechanism for hypernatremia rather than simple dehydration.

While it might not be that electrolyte imbalances aggravate the severity of COVID-19 in a patient, it may be that the underlying severe disease is resulting in physiologic derangements that manifest partly as electrolyte imbalances. These findings suggest the possible role of hyponatremia, hypocalcemia and hypernatremia in the risk stratification, prognostication and clinical decision-making when treating COVID-19 patients. As conventional biomarkers like IL-6 and CRP are expensive to test especially in rural healthcare centres, the measurement of electrolytes can serve as an additional tool to triage scarce healthcare resources such as ICU beds and ventilators due to its ease of access. Hypernatremia may be a clinically useful indicator of progression to a poor outcome due to its high LR+ of 4.0 resulting in a PPV of 55% in the general population of COVID patients.

The strengths of our study lie in the large number of studies analyzed looking at a broad range of electrolyte imbalances. None of our included studies had a high risk-of-bias according to the NOS scale, increasing the quality of findings. We employed a rigorous methodology according to international guidelines pre-specified in our protocol. Additionally, we pooled maximally adjusted estimates to account for potential confounders and assessed the prognostic value of each electrolyte imbalance by calculating their overall sensitivity, specificity, positive likelihood ratio, negative likelihood ratio and area the under curve scores. Our findings were also robust to pre-specified subgroup and sensitivity analyses.

### Limitations

Firstly, there were insufficient studies looking at the same outcome and same electrolyte imbalance for a statistically-powered meta-regression and funnel plot for the assessment of publication bias. We were unable to conduct some meaningful subgroup analyses to explain heterogeneity, but this potentially could be explained by the differing impacts of the pandemic on different healthcare systems globally, as well as their varying management strategies. This may be confirmed using future studies for subgroup analyses stratified by country or region. Secondly, participants may be admitted with differing COVID-19 baseline severities, with some being admitted directly to the ICU with an already severe infection. We are unable to discern this group from those that deteriorated after admission, potentially introducing a source of bias as participants who are severely infected will be more prone to having a poor outcome. Nonetheless, we mitigated this by marking down the study’s representativeness on the NOS scale. Thirdly, our results do not allow us to interpret the causality of the association as it is unclear whether the electrolyte imbalances further aggravate participants with COVID-19 or whether its just a general indication of poor health. Furthermore, the temporal sequence between COVID-19 diagnosis and the presence of electrolyte imbalances is hard to establish. Studies also did not monitor the progression of these imbalances throughout the length of hospital stay. Additionally, we acknowledge that respiratory infections such as COVID-19 commonly result in dehydration because of pyrexia or tachypnaea and that our association could be confounded by abnormalities such as blood volume and osmolarity. Not all our studies assessed the specific etiology of the electrolyte imbalance, whether it is hypovolemic, euvolemic or hypervolemic which could potentially influence management.

## CONCLUSION

In this multi-adjusted observational meta-analysis of 26,897 participants with COVID-19, hyponatremia, hypernatremia and hypocalcemia were associated with a 2-fold, 4-fold and 3-fold increased odds of poor clinical outcome respectively. We also observed that hypernatremia had a specificity of 97% independent of CRP and IL-6 levels, highlighting its potential use as a unique clinical indicator of poor disease progression. Our findings are pertinent to triaging and risk assessment of COVID-19 patients, especially since severe COVID-19 patients continue to take up significant healthcare resources. Future interventional studies and randomized controlled trials should look at whether correcting for these electrolyte imbalances via resuscitation strategies, fluid replacement or supplements can mitigate the odds of poor outcome.

## Data Availability

All data produced in the present study are available upon reasonable request to the authors

## ACKNOWLEDGEMENTS

We thank Dr Chan Yong Hiuak (Founding Mentor, NUS Medicine Biostatistics Unit) for his statistical review of our manuscript.

