## Supplemental Material for "Associations and prognostic accuracy of electrolyte imbalances in predicting poor COVID-19 outcome: a systematic review and meta-analysis"

### SUPPLEMENTAL METHODS

#### Search Strategy

1 “hypernatremia” OR “hyponatremia” OR “dysnatremia” OR “hyperchloremia” OR “hypochloremia” OR “dyschloremia” OR “hyperkalemia” OR “hypokalemia” OR “dyskalemia” OR “hypermagnesemia” OR “hypomagnesemia” OR “dysmagnesemia” OR “hyperphosphatemia” OR “hypophosphatemia” OR “dysphosphatemia” OR “hypercalcemia” OR “hypocalcemia” OR “dyscalcemia”
2 (“abnormal” OR “dysfunction” OR “disorder” OR “altered” OR “alterations” OR “irregularity” OR “imbalance”) AND (“electrolyte” OR “sodium” OR “potassium” OR “chloride” OR “magnesium” OR “phosphate” OR “calcium”)

3	1 OR 2

4	“COVID-19” OR “SARS-CoV-2” OR “sars cov 2” OR “sars cov2” OR “covid-19” OR “covid 19” OR “covid19” OR “covid 2019” OR “coronavirus disease 2019” OR “coronavirus disease 19” OR “severe acute respiratory syndrome coronavirus 2” OR “2019ncov” OR “SARS2” OR “SARS-CoV-19” OR “novel cov” OR “cov22”

5	3 AND 4

#### Study Selection, Data Extraction, Risk of Bias Assessment and Quality of Evidence

We extracted key data including the first author, year published, study design, country, sample size, percentage male, mean/median age, type of electrolyte imbalances studied and its cut-off values, outcomes, statistical methods and other key findings. We extracted key data and assessed the risk of bias using the Newcastle-Ottawa Scale, which grades studies as having a high (<5 stars), moderate (5-7 stars) or low (≥8 stars) risk of bias.[1]

#### Statistical Analyses

We used an inverse variance-weighted mixed-effects model, which consists of both a fixed-effects model and random effects model to pool subgroup and study-level estimates respectively. If studies stratified their population into subgroups (e.g. by severity of electrolyte imbalance), we pooled subgroup-level estimates using a fixed-effects model to derive a study-level estimate. For dichotomous clinical outcomes, given that hazard and odds ratios numerically approximate one another when follow-up duration, average rate of event and magnitude of risk is low,[2] we pooled maximally adjusted odds ratios and hazard ratios together as an overall odds ratio if the above criteria were met. As not all studies computed an adjusted summary estimate, we also pooled raw frequency counts for each dichotomous outcome via the Mantel-Haenszel method. For continuous outcomes including laboratory parameters and hospitalization time, we pooled mean differences using an inverse variance-weighted random-effects model. If the study did not report means and standard deviations, we estimated them from the reported medians, interquartile ranges and sample sizes using a method outlined by Luo et al.[3] and Hozo et al.[4] We defined poor outcome as a composite of mortality, ICU admission, respiratory support and ARDS due to their resource-intensive nature, in line with previous landmark studies on severe COVID.[5, 6] If studies reported more than two of the aforementioned outcomes (e.g. both mortality and respiratory support), we selected the outcome with greater raw frequency counts to prevent double-counting of study participants. To investigate potential sources of heterogeneity, we pre-specified various study-level characteristics to perform subgroup or sensitivity analyses for the following characteristics: (1) time point of measurement of electrolyte imbalance, (2) age, (3) gender, (4) covariates adjusted, (5) definition of electrolyte imbalance, (6) setting/country, (7) study design, (8) risk of bias.

### SUPPLEMENTAL FIGURES

Supplemental Figure S1**: Forest plot showing the pooled unadjusted odds ratios (a) and adjusted odds ratios (b) of the association between electrolyte imbalances and mortality, stratified by the type of electrolyte imbalance**

Legend: Black diamonds are the estimated pooled odds ratios for each random-effects meta-analysis; blue/red boxes reflect the relative weight apportioned to studies in the meta-analysis

Supplemental Figure S1a: Mortality, unadjusted odds ratios, stratified by the type of electrolyte imbalance

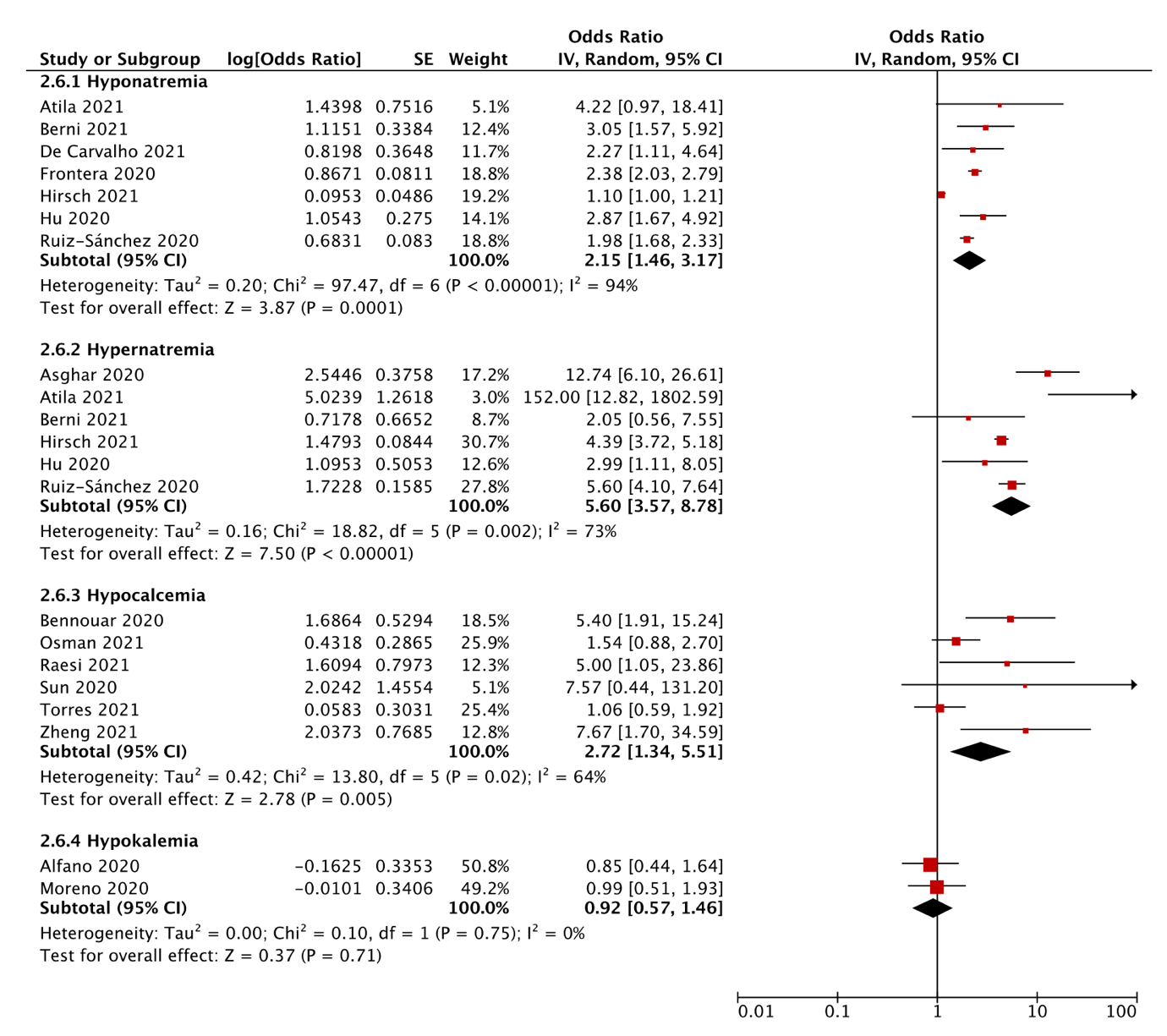

Supplemental Figure S1b: Mortality, adjusted odds ratios, stratified by the type of electrolyte imbalance

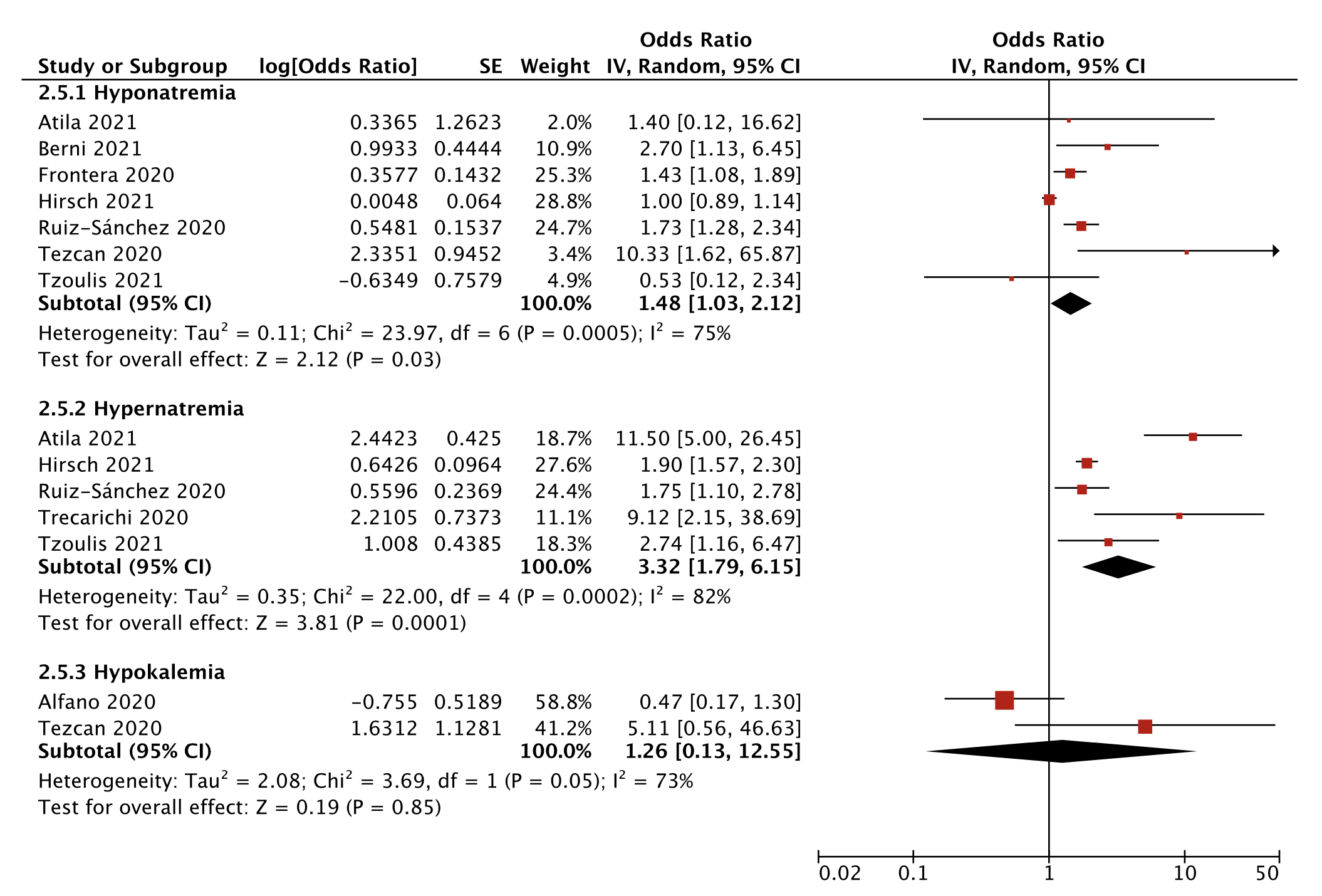

Supplemental Figure S2**: Forest plot showing the unadjusted association between the severity of hyponatremia and mortality**

Legend: Black diamonds are the estimated pooled odds ratios for each random-effects meta-analysis; blue boxes reflect the relative weight apportioned to studies in the meta-analysis

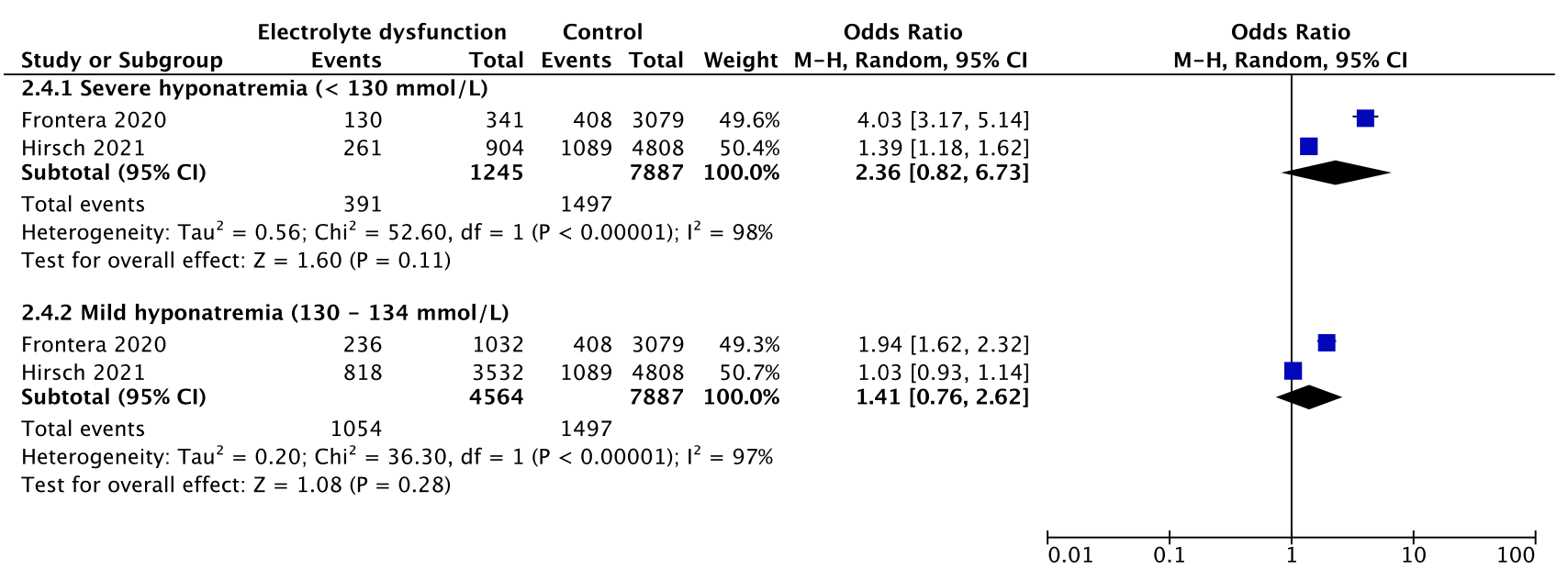

Supplemental Figure S3**: Forest plot showing the unadjusted association between electrolyte imbalances with ICU admission, stratified by the type of electrolyte imbalance**

Legend: Black diamonds are the estimated pooled odds ratios for each random-effects meta-analysis; blue boxes reflect the relative weight apportioned to studies in the meta-analysis

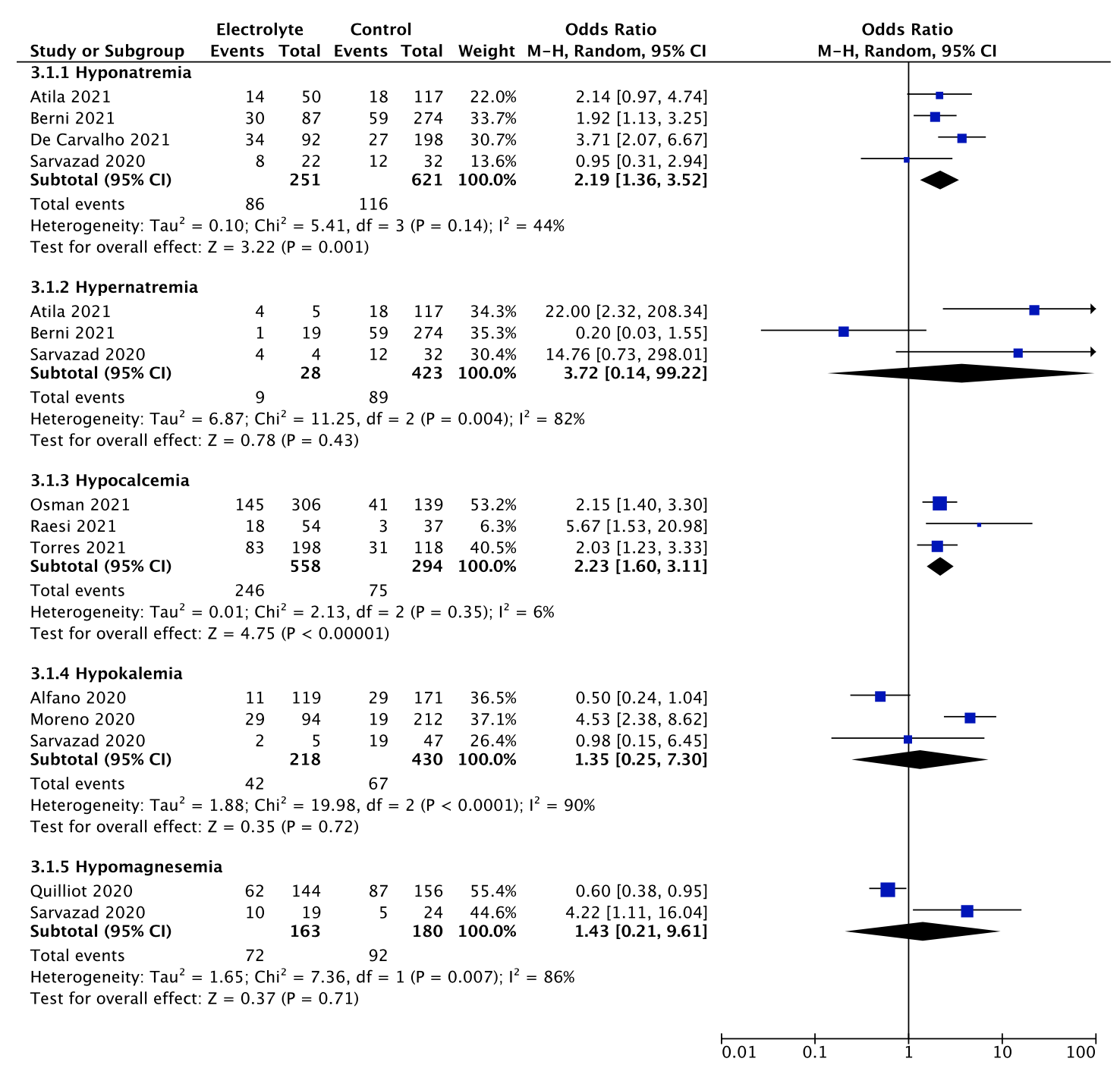

Supplemental Figure S4**: Forest plot showing the (a) unadjusted and (b) adjusted association between electrolyte imbalances and respiratory support, stratified by the type of electrolyte imbalance**

Legend: Black diamonds are the estimated pooled odds ratios for each random-effects meta-analysis; red boxes reflect the relative weight apportioned to studies in the meta-analysis

Supplemental Figure S4a: Respiratory support, unadjusted association, stratified by the type of electrolyte imbalance

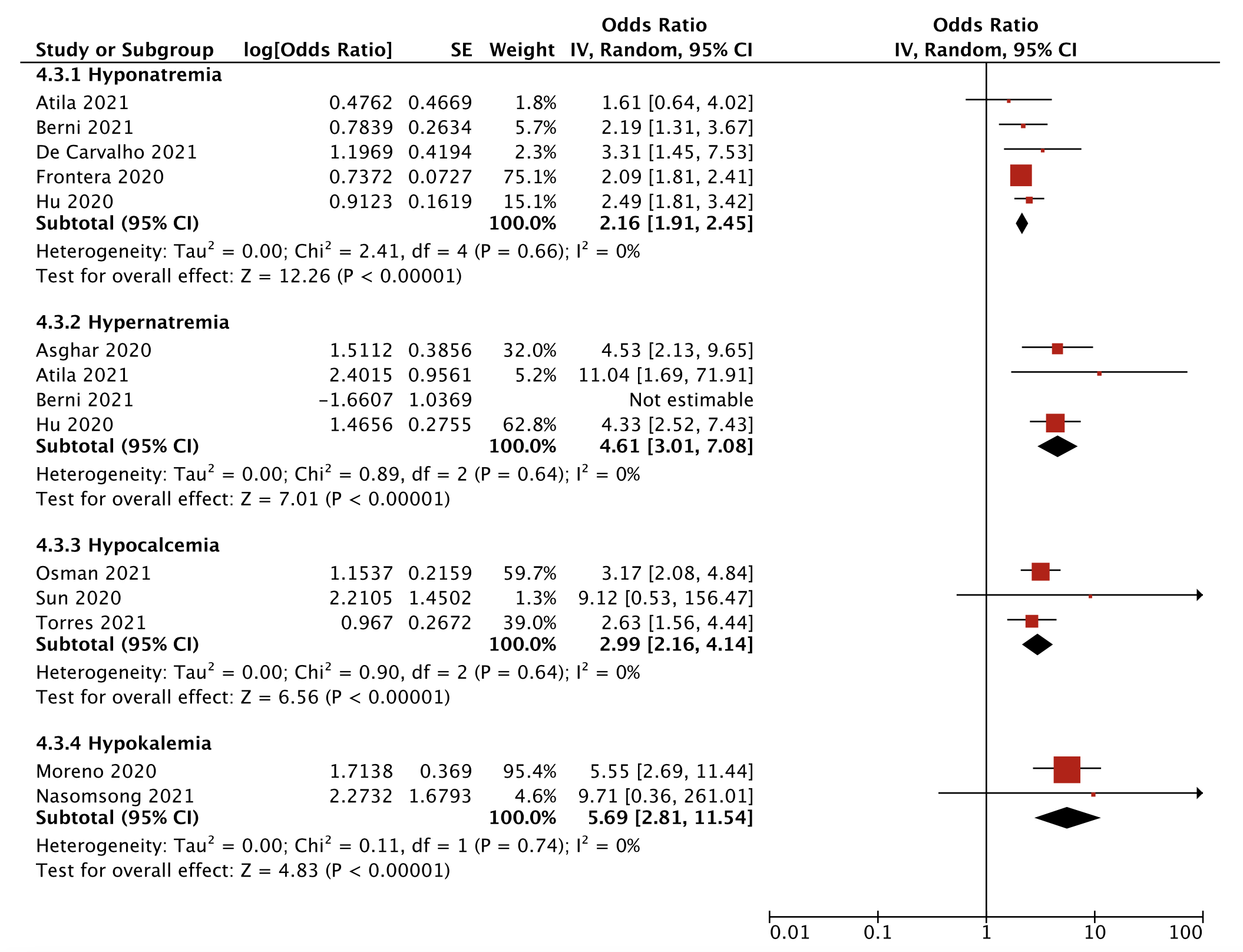

Supplemental Figure S4b: Respiratory support, adjusted association

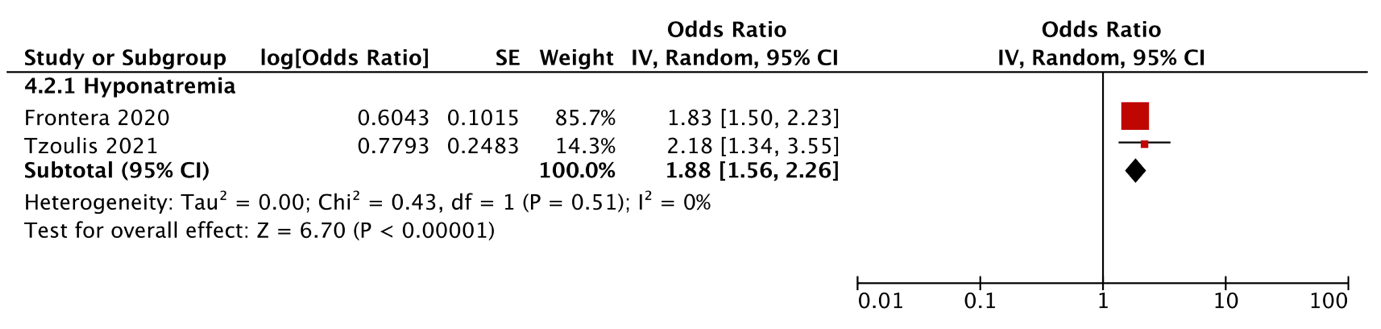

Supplemental Figure S5**: Forest plot showing the unadjusted association of (a) hyponatremia and (b) hypernatremia with respiratory support, stratified by the type of ventilation**

Legend: Black diamonds are the estimated pooled odds ratios for each random-effects meta-analysis; blue boxes reflect the relative weight apportioned to studies in the meta-analysis

Supplemental Figure S5a: Hyponatremia, respiratory support, stratified by type of ventilation

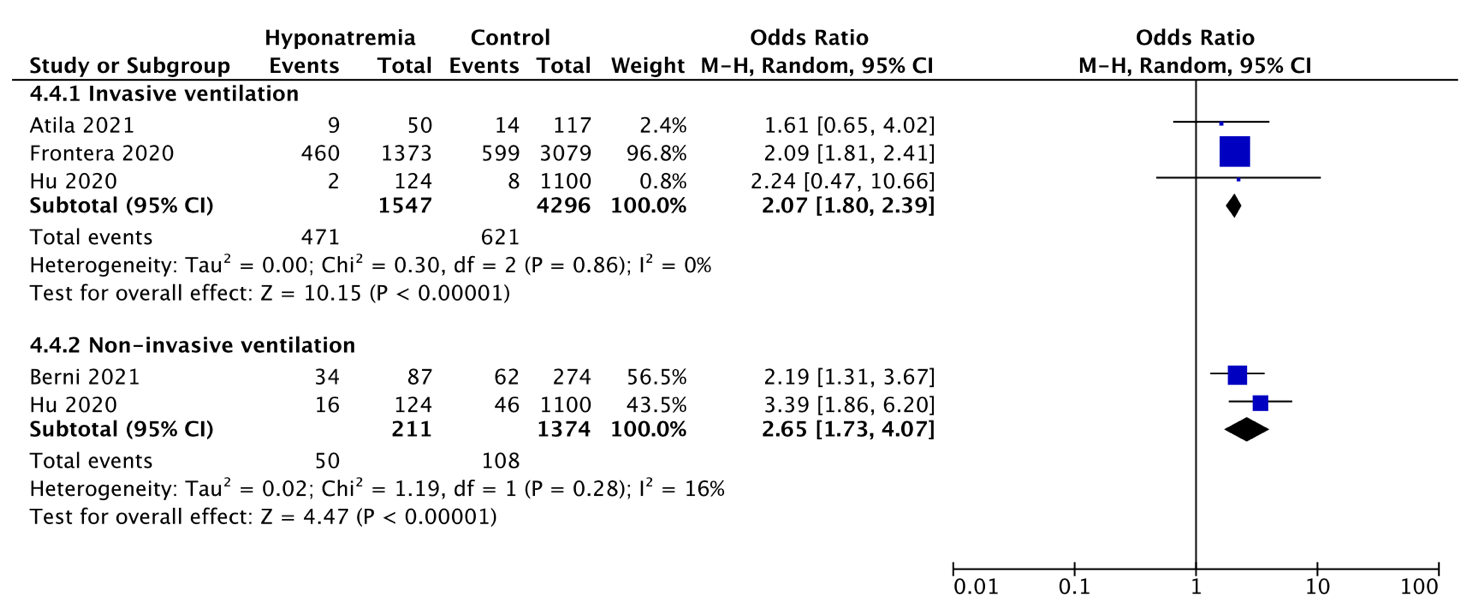

Supplemental Figure S5b: Hypernatremia, respiratory support, stratified by type of ventilation

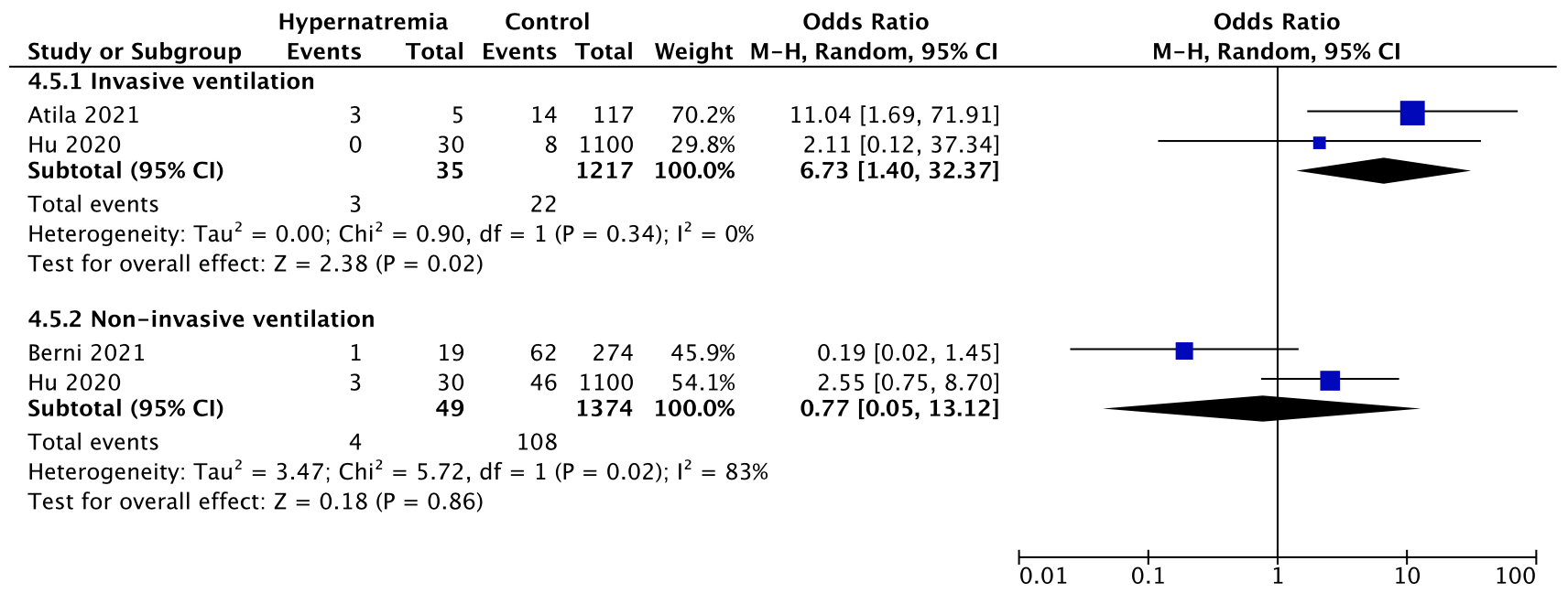

Supplemental Figure S6**: Forest plot showing the unadjusted association between electrolyte imbalances and acute respiratory distress syndrome**

Legend: Black diamonds are the estimated pooled odds ratios for each random-effects meta-analysis; blue boxes reflect the relative weight apportioned to studies in the meta-analysis

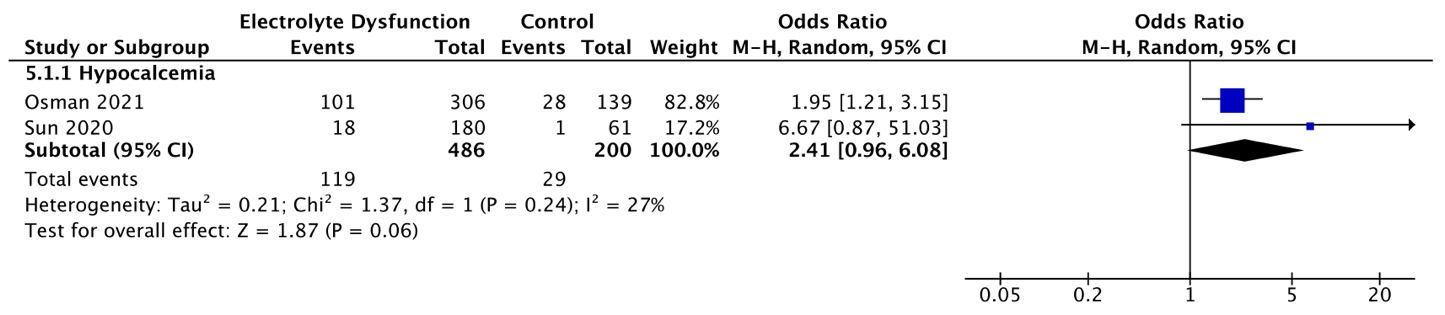

Supplemental Figure S7**: Forest plot showing the unadjusted association between electrolyte imbalances and acute kidney injury**

Legend: Black diamonds are the estimated pooled odds ratios for each random-effects meta-analysis; blue boxes reflect the relative weight apportioned to studies in the meta-analysis

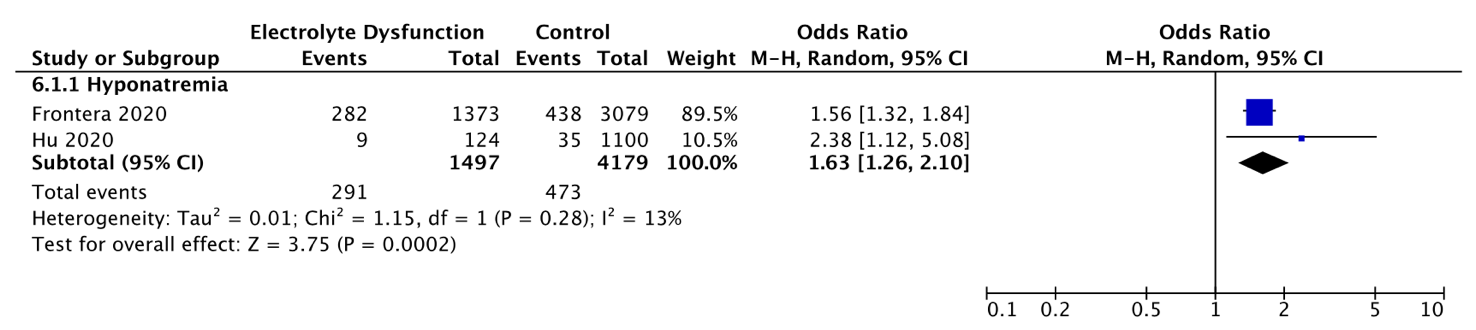

Supplemental Figure S8**: Forest plot showing the mean differences for (a) serum creatinine, (b) C-reactive protein and (c) hospitalization time, stratified by the type of electrolyte imbalance.**

Legend: Black diamonds are the estimated pooled mean differences for each random-effects meta-analysis; green boxes reflect the relative weight apportioned to studies in the meta-analysis

Supplemental Figure S8a: Serum creatinine (mg/dL), stratified by the type of electrolyte imbalance

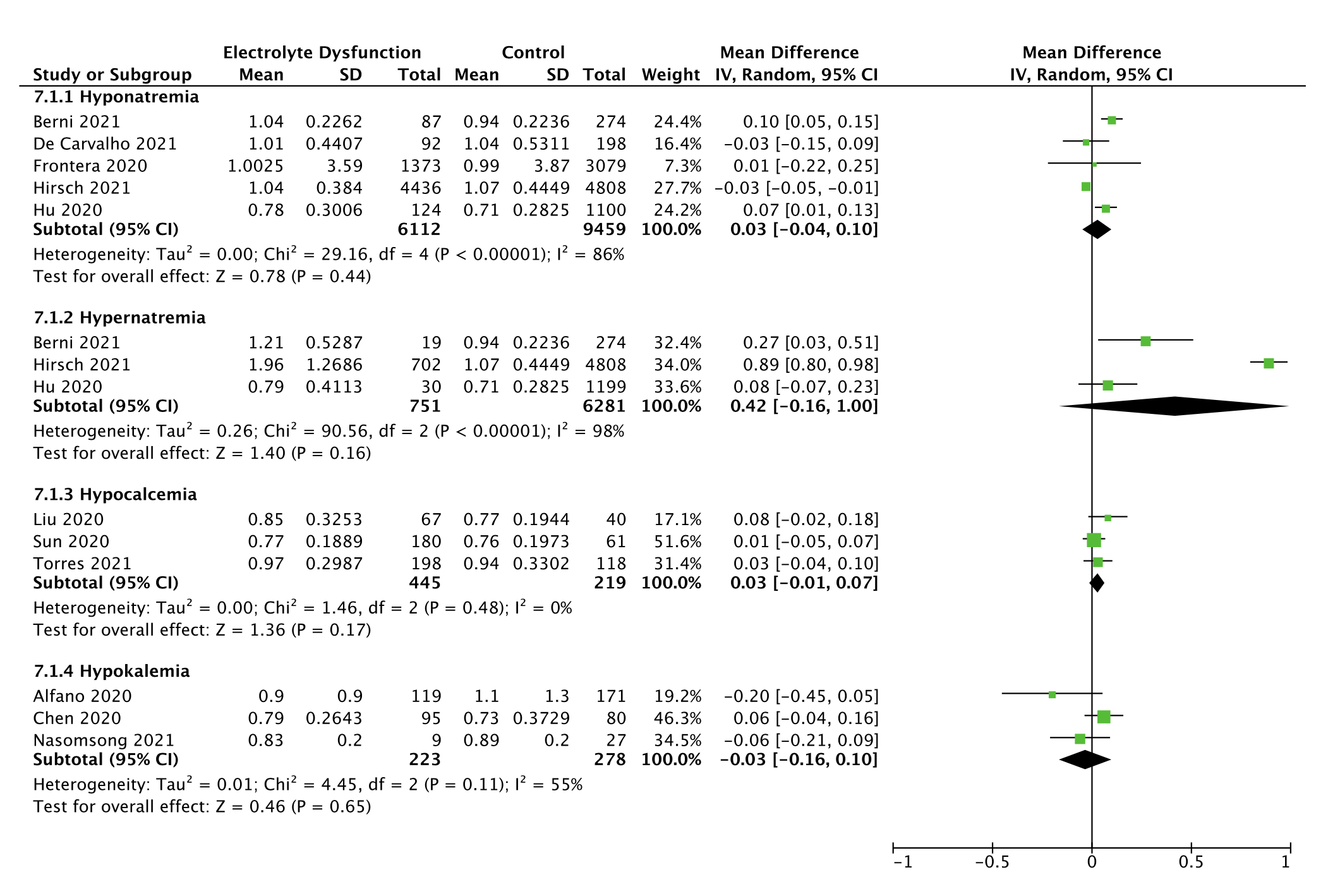

Supplemental Figure S8b: C-reactive protein (mg/L), stratified by type of electrolyte imbalance

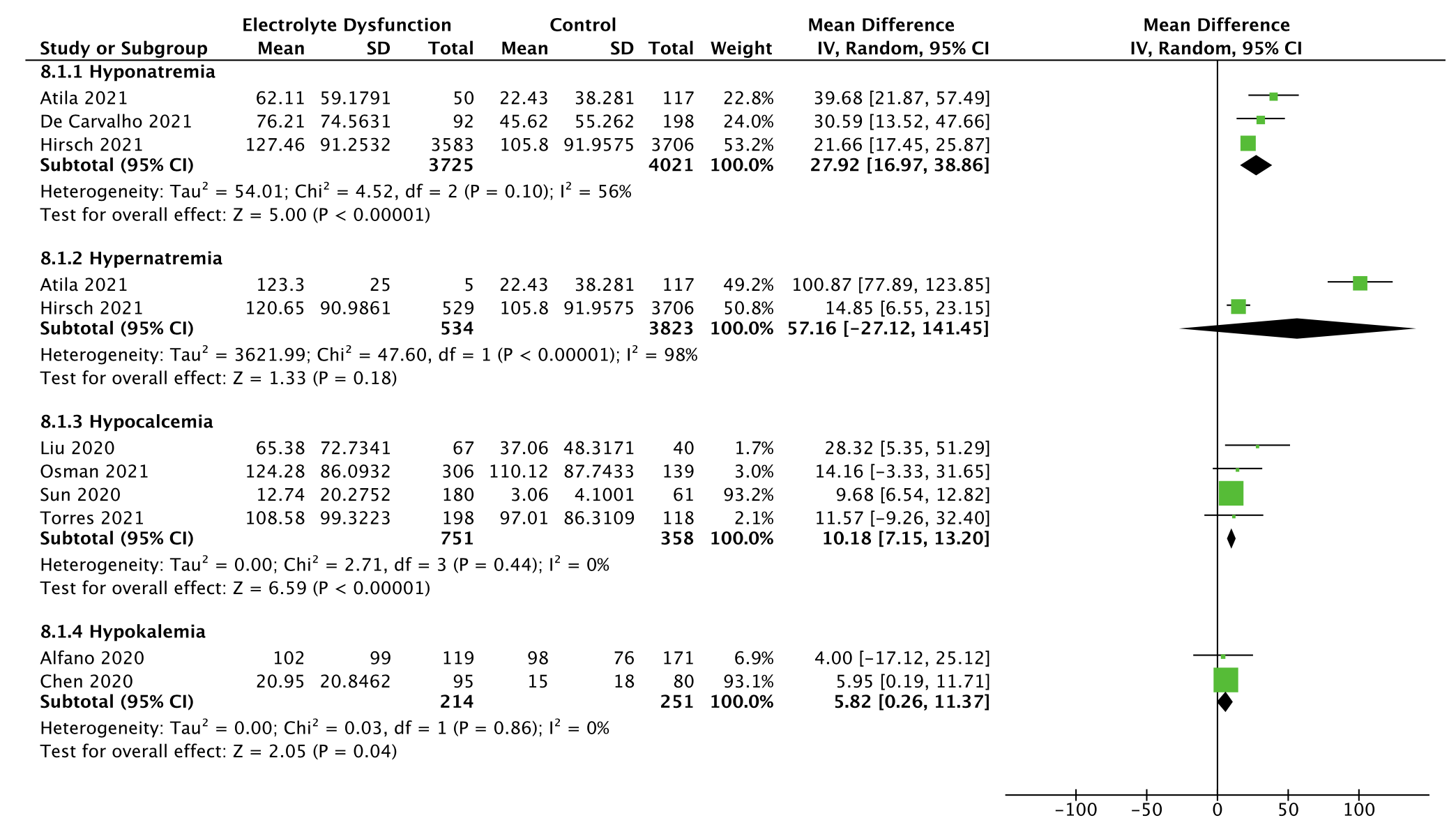

Supplemental Figure S8c: Hospitalization time (days), stratified by type of electrolyte imbalance

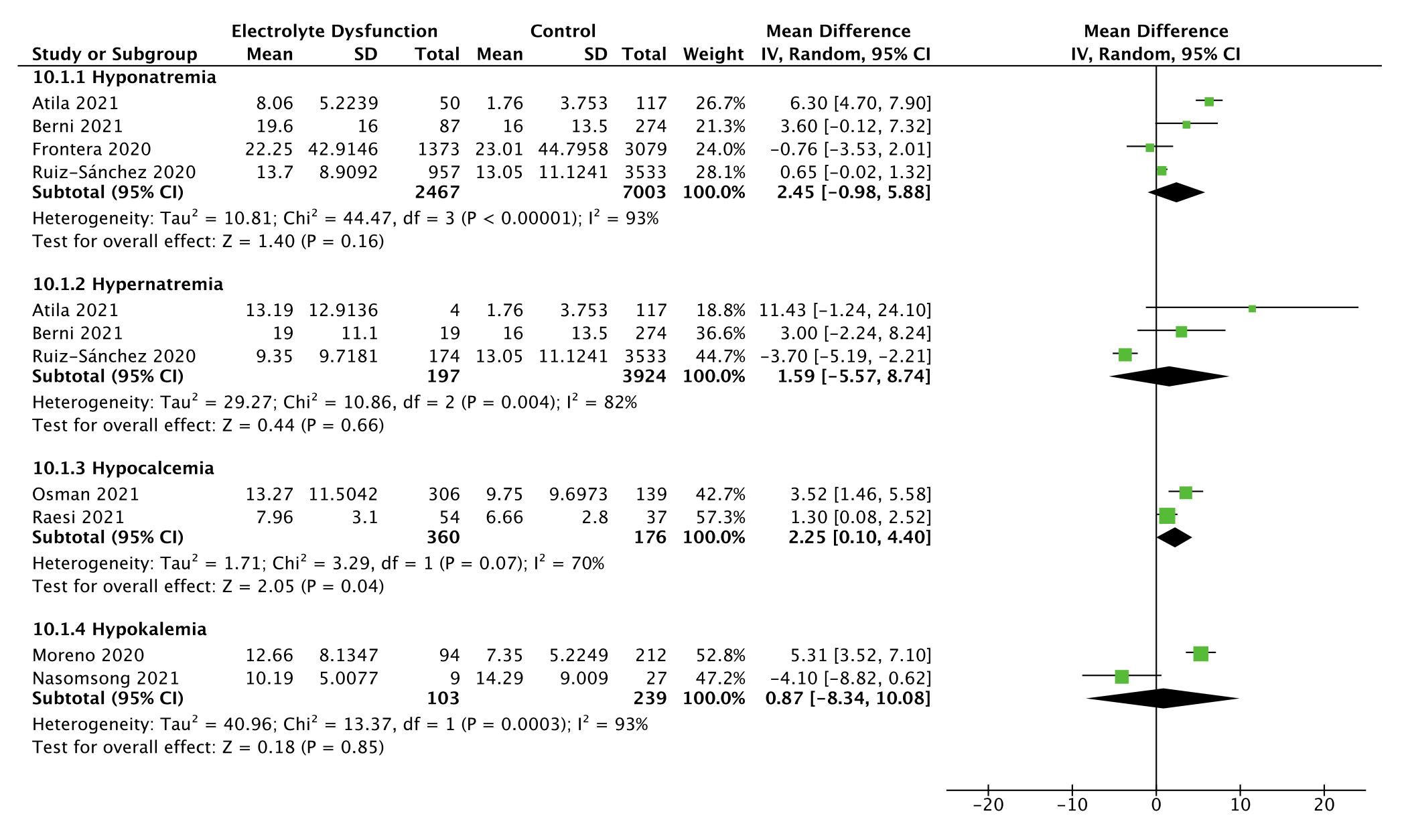

Supplemental Figure S9**: Summary receiver operator characteristic curve of (a) hyponatremia and (b) hypokalemia**

Supplemental Figure S9a: Summary ROC curve of hyponatremia and poor outcome

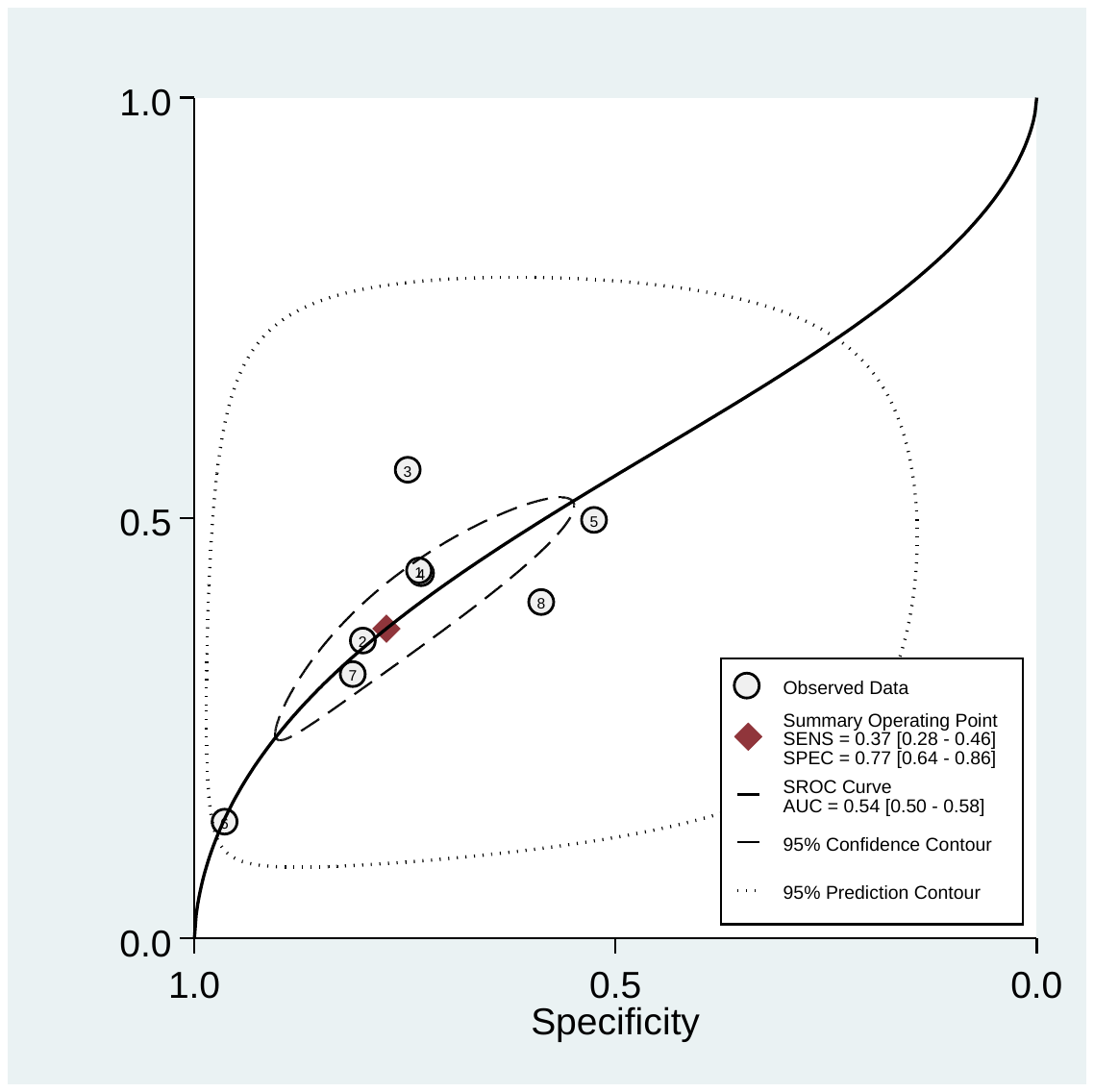

Supplemental Figure S9b: Summary ROC curve of hypokalemia and poor outcome

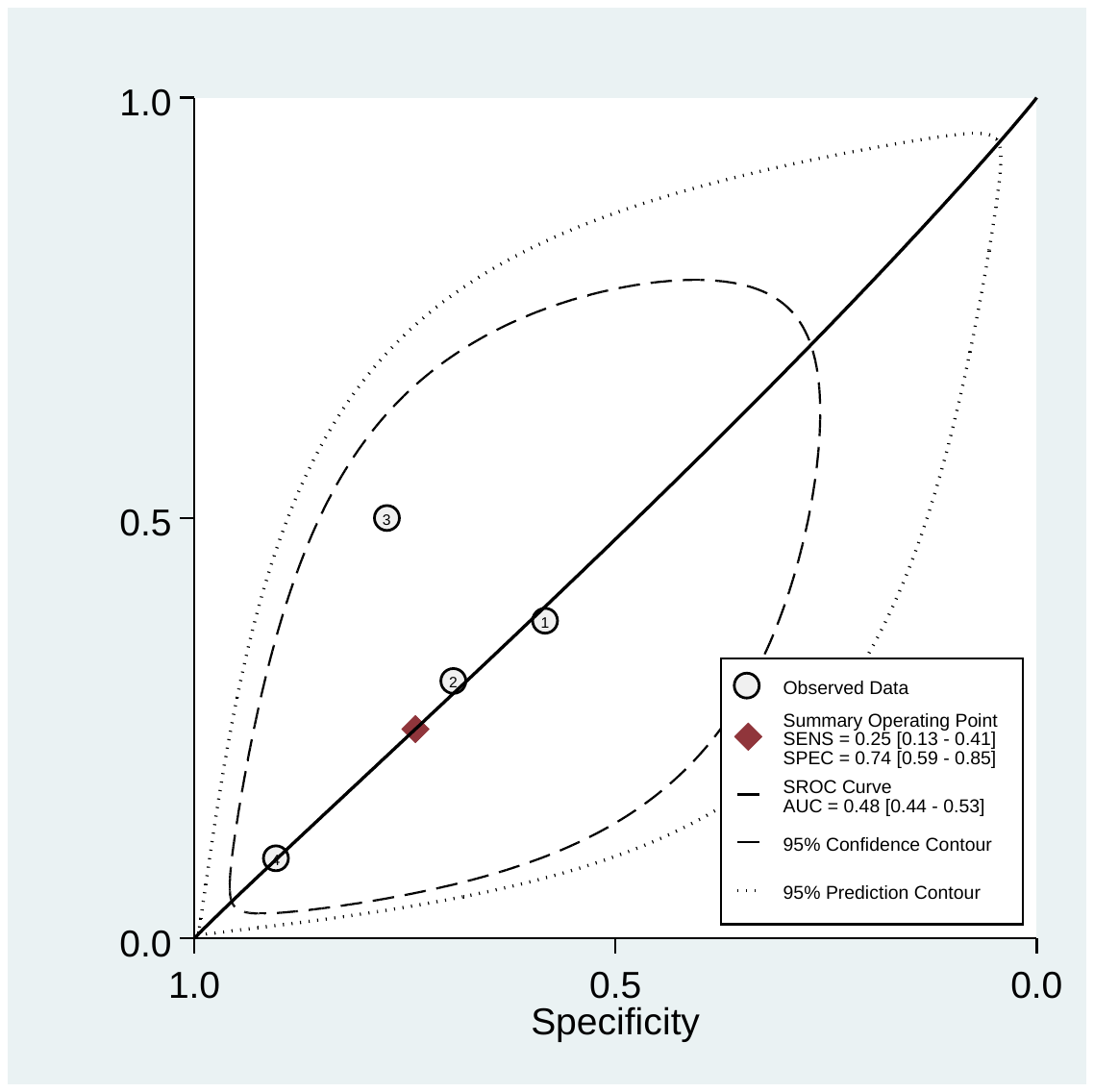

### SUPPLEMENTAL TABLES

Supplemental Table S1**: PRISMA Checklist**

| **Section and Topic** | **Item #** | **Checklist item** | **Page number where item is reported** |
| --- | --- | --- | --- |
| **TITLE** | | |  |
| Title | 1 | Identify the report as a systematic review. | 1 |
| **ABSTRACT** | | |  |
| Abstract | 2 | See the PRISMA 2020 for Abstracts checklist. | 3, 4 |
| **INTRODUCTION** | | |  |
| Rationale | 3 | Describe the rationale for the review in the context of existing knowledge. | 5 |
| Objectives | 4 | Provide an explicit statement of the objective(s) or question(s) the review addresses. | 5, 6 |
| **METHODS** | | |  |
| Eligibility criteria | 5 | Specify the inclusion and exclusion criteria for the review and how studies were grouped for the syntheses. | 6, 7 |
| Information sources | 6 | Specify all databases, registers, websites, organisations, reference lists and other sources searched or consulted to identify studies. Specify the date when each source was last searched or consulted. | 6 |
| Search strategy | 7 | Present the full search strategies for all databases, registers and websites, including any filters and limits used. | Supplement |
| Selection process | 8 | Specify the methods used to decide whether a study met the inclusion criteria of the review, including how many reviewers screened each record and each report retrieved, whether they worked independently, and if applicable, details of automation tools used in the process. | 6, 7 |
| Data collection process | 9 | Specify the methods used to collect data from reports, including how many reviewers collected data from each report, whether they worked independently, any processes for obtaining or confirming data from study investigators, and if applicable, details of automation tools used in the process. | 6, 7, Supplement |
| Data items | 10a | List and define all outcomes for which data were sought. Specify whether all results that were compatible with each outcome domain in each study were sought (e.g. for all measures, time points, analyses), and if not, the methods used to decide which results to collect. | 6, 7, Supplement |
|  | 10b | List and define all other variables for which data were sought (e.g. participant and intervention characteristics, funding sources). Describe any assumptions made about any missing or unclear information. | 6, 7, Supplement |
| Study risk of bias assessment | 11 | Specify the methods used to assess risk of bias in the included studies, including details of the tool(s) used, how many reviewers assessed each study and whether they worked independently, and if applicable, details of automation tools used in the process. | 6, 7 |
| Effect measures | 12 | Specify for each outcome the effect measure(s) (e.g. risk ratio, mean difference) used in the synthesis or presentation of results. | 7, 8 |
| Synthesis methods | 13a | Describe the processes used to decide which studies were eligible for each synthesis (e.g. tabulating the study intervention characteristics and comparing against the planned groups for each synthesis (item #5)). | 7, 8, Supplement |
|  | 13b | Describe any methods required to prepare the data for presentation or synthesis, such as handling of missing summary statistics, or data conversions. | 7, 8, Supplement |
|  | 13c | Describe any methods used to tabulate or visually display results of individual studies and syntheses. | 7, 8 |
|  | 13d | Describe any methods used to synthesize results and provide a rationale for the choice(s). If meta-analysis was performed, describe the model(s), method(s) to identify the presence and extent of statistical heterogeneity, and software package(s) used. | 7, 8, Supplement |
|  | 13e | Describe any methods used to explore possible causes of heterogeneity among study results (e.g. subgroup analysis, meta-regression). | 7, 8 |
|  | 13f | Describe any sensitivity analyses conducted to assess robustness of the synthesized results. | 7, 8 |
| Reporting bias assessment | 14 | Describe any methods used to assess risk of bias due to missing results in a synthesis (arising from reporting biases). | 6,7 |
| Certainty assessment | 15 | Describe any methods used to assess certainty (or confidence) in the body of evidence for an outcome. | 6, 7 |
| **RESULTS** | | |  |
| Study selection | 16a | Describe the results of the search and selection process, from the number of records identified in the search to the number of studies included in the review, ideally using a flow diagram. | 8 |
|  | 16b | Cite studies that might appear to meet the inclusion criteria, but which were excluded, and explain why they were excluded. | 8, Figure 1 |
| Study characteristics | 17 | Cite each included study and present its characteristics. | 8 |
| Risk of bias in studies | 18 | Present assessments of risk of bias for each included study. | 8 |
| Results of individual studies | 19 | For all outcomes, present, for each study: (a) summary statistics for each group (where appropriate) and (b) an effect estimate and its precision (e.g. confidence/credible interval), ideally using structured tables or plots. | 10-15, Supplement |

| Results of syntheses | 20a | For each synthesis, briefly summarise the characteristics and risk of bias among contributing studies. | 10-15, Supplement |
| --- | --- | --- | --- |
|  | 20b | Present results of all statistical syntheses conducted. If meta-analysis was done, present for each the summary estimate and its precision (e.g. confidence/credible interval) and measures of statistical heterogeneity. If comparing groups, describe the direction of the effect. | 10-15, Supplement |
|  | 20c | Present results of all investigations of possible causes of heterogeneity among study results. | 10-15, Supplement |
|  | 20d | Present results of all sensitivity analyses conducted to assess the robustness of the synthesized results. | 10-15, Supplement |
| Reporting biases | 21 | Present assessments of risk of bias due to missing results (arising from reporting biases) for each synthesis assessed. | 10-15, Supplement |
| Certainty of evidence | 22 | Present assessments of certainty (or confidence) in the body of evidence for each outcome assessed. | 8 |
| **DISCUSSION** | | |  |
| Discussion | 23a | Provide a general interpretation of the results in the context of other evidence. | 16, 17 |
|  | 23b | Discuss any limitations of the evidence included in the review. | 19 |
|  | 23c | Discuss any limitations of the review processes used. | 19 |
|  | 23d | Discuss implications of the results for practice, policy, and future research. | 20 |
| **OTHER INFORMATION** | | |  |
| Registration and protocol | 24a | Provide registration information for the review, including register name and registration number, or state that the review was not registered. | 6 |
|  | 24b | Indicate where the review protocol can be accessed, or state that a protocol was not prepared. | 6 |
|  | 24c | Describe and explain any amendments to information provided at registration or in the protocol. | - |
| Support | 25 | Describe sources of financial or non-financial support for the review, and the role of the funders or sponsors in the review. | 1 |
| Competing interests | 26 | Declare any competing interests of review authors. | 21 |
| Availability of data, code and other materials | 27 | Report which of the following are publicly available and where they can be found: template data collection forms; data extracted from included studies; data used for all analyses; analytic code; any other materials used in the review. | 1 |

Supplemental Table S2**: Risk-of-bias assessment of cohort and cross-sectional studies using the Newcastle-Ottawa Scale**

| **Study** | **Representativeness of participants with electrolyte imbalance** | **Non-exposed group drawn from same community as exposed group** | **Exposure ascertained through lab tests** | **Outcome of interest was not present at start of study** | **Adjusts for age and sex** | **Adjusts for ≥ 3 other covariates** | **Assessment of outcome** | **Median follow-up at least 2 weeks** | **Adequacy of follow-up** | **Total/9** | **Risk of bias*** |
| --- | --- | --- | --- | --- | --- | --- | --- | --- | --- | --- | --- |
| Alfano, 2020 | 1 | 1 | 1 | 1 | 1 | 0 | 1 | 0 | 1 | 7 | Moderate |
| Asghar, 2020 | 1 | 1 | 1 | 1 | 0 | 0 | 1 | 1 | 1 | 7 | Moderate |
| Atila, 2021 | 1 | 1 | 1 | 1 | 1 | 1 | 1 | 1 | 1 | 9 | Low |
| Bennouar, 2020 | 1 | 1 | 1 | 0 | 1 | 1 | 1 | 0 | 1 | 7 | Moderate |
| Berni, 2021 | 1 | 1 | 1 | 1 | 1 | 0 | 1 | 1 | 1 | 8 | Low |
| Carvalho, 2021 | 1 | 1 | 1 | 1 | 1 | 1 | 1 | 0 | 0 | 7 | Moderate |
| Chen, 2020 | 1 | 1 | 1 | 1 | 0 | 0 | 1 | 0 | 1 | 6 | Moderate |
| Frontera, 2020 | 1 | 1 | 1 | 1 | 1 | 1 | 1 | 0 | 1 | 8 | Low |
| Hirsch, 2021 | 1 | 1 | 1 | 1 | 1 | 1 | 1 | 1 | 0 | 8 | Low |
| Hu, 2020 | 1 | 1 | 1 | 0 | 0 | 0 | 1 | 0 | 1 | 5 | Moderate |
| Hu, 2021 | 1 | 1 | 1 | 1 | 0 | 0 | 1 | 0 | 1 | 6 | Moderate |
| Liu, 2020 | 1 | 1 | 1 | 0 | 1 | 0 | 1 | 1 | 1 | 7 | Moderate |
| Ma, 2020 | 1 | 1 | 1 | 1 | 1 | 0 | 1 | 1 | 1 | 8 | Low |
| Moreno, 2020 | 1 | 1 | 1 | 1 | 1 | 1 | 1 | 1 | 1 | 9 | Low |
| Nasomsong, 2021 | 1 | 1 | 1 | 0 | 0 | 0 | 1 | 0 | 1 | 5 | Moderate |
| Osman, 2021 | 1 | 1 | 1 | 0 | 0 | 0 | 1 | 0 | 1 | 5 | Moderate |
| Quilliot, 2020 | 1 | 1 | 1 | 0 | 0 | 0 | 1 | 0 | 1 | 5 | Moderate |
| Raesi, 2021 | 1 | 1 | 1 | 0 | 0 | 0 | 1 | 0 | 1 | 5 | Moderate |
| Sanchez, 2020 | 1 | 1 | 1 | 1 | 1 | 1 | 1 | 1 | 1 | 9 | Low |
| Sarvazad, 2020 | 1 | 1 | 1 | 0 | 0 | 0 | 1 | 0 | 1 | 5 | Moderate |
| Sun, 2020 | 1 | 1 | 1 | 0 | 0 | 0 | 1 | 1 | 1 | 6 | Moderate |
| Tezcan, 2020 | 1 | 1 | 1 | 1 | 1 | 1 | 1 | 0 | 1 | 8 | Low |
| Torres, 2021 | 1 | 1 | 1 | 1 | 0 | 1 | 1 | 0 | 1 | 7 | Moderate |
| Trecarichi, 2020 | 1 | 1 | 1 | 1 | 0 | 0 | 1 | 0 | 1 | 6 | Moderate |
| Tzoulis, 2021 | 1 | 1 | 1 | 1 | 1 | 1 | 1 | 0 | 1 | 8 | Low |
| Wu, 2020 | 1 | 1 | 1 | 1 | 0 | 1 | 1 | 0 | 1 | 7 | Moderate |
| Zheng, 2021 | 0 | 1 | 1 | 1 | 0 | 0 | 1 | 0 | 1 | 5 | Moderate |
| Zhou, 2020 | 0 | 1 | 1 | 1 | 0 | 0 | 1 | 0 | 1 | 5 | Moderate |

*8-9 stars: Low, 5-7 stars: Moderate, 0-4 stars: High

Supplemental Table S3**: Evaluation of quality of pooled evidence assessing causal effect using the Grading of Recommendations Assessment, Development and Evaluation (GRADE) framework**

| Outcomes | Pooled outcomes (95% CI) | No. of participants (no. of included studies) | Statistical heterogeneity | Risk of bias | Imprecision | Inconsistency | Indirectness | Publication bias | Quality of evidence (GRADE) |
| --- | --- | --- | --- | --- | --- | --- | --- | --- | --- |
| Poor outcome in hyponatremia | 1.65 (1.09-2.51) | 19,951 (5 studies) | I^2^=91% (p<0.001) | Not serious | Not serious | Serious^2^ | Not serious | Not serious | Low^3^ |
| Poor outcome in hypernatremia | 2.10 (1.80-2.44) | 14,898 (3 studies) | I^2^=0%  (p=0.39) | Not serious | Not serious | Not serious | Not serious | Not serious | Moderate^3^ |
| Poor outcome in hypocalcemia | 3.31 (2.24-4.88) | 1,374 (6 studies) | I^2^=25% (p=0.25) | Serious^4^ | Not serious | Not serious | Not serious | Not serious | Low^3^ |
| Poor outcome in hypokalemia | 1.26 (0.13-12.55) | 728 (2 studies) | I^2^=73%  (p=0.05) | Not serious | Serious^1^ | Serious^2^ | Not serious | Not serious | Very low |
| Poor outcome in hypomagnesemia | 1.43 (0.21-9.60) | 358 (2 studies) | I^2^=86% (p<0.05) | Serious^4^ | Serious^1^ | Serious^2^ | Not serious | Not serious | Very low |

Quality of evidence for observational studies is graded starting at low quality for a causal effect, and downgraded or upgraded based on the following criteria: risk of bias, imprecision, inconsistency, indirectness and publication bias. 1. Downgraded by one level as only two studies were pooled. 2. Downgraded by one level due to significant heterogeneity detected. 3. Upgraded by one level due to large magnitude of effect. 4. Associations were unadjusted

Supplemental Table S4**: Evaluation of quality of pooled evidence assessing prognostic accuracy using the Grading of Recommendations Assessment, Development and Evaluation (GRADE) framework**

| Outcomes | Sensitivity (95% CI)  Statistical heterogeneity | Specificity (95%CI)  Statistical heterogeneity | No. of participants (no. of included studies) | Risk of bias | Imprecision | Inconsistency | Indirectness | Publication bias | Quality of evidence (GRADE) |
| --- | --- | --- | --- | --- | --- | --- | --- | --- | --- |
| Prognostic accuracy of hyponatremia | 0.37 (0.28-0.46)  I^2^=98% | 0.77 (0.64-0.86)  I^2^=99% | 20,282 (8 studies) | Not serious | Not serious | Serious^2^ | Not serious | Not serious | Moderate |
| Prognostic accuracy of hypernatremia | 0.13 (0.07-0.22)  I^2^=97% | 0.97 (0.94-0.98)  I^2^=95% | 10,798 (6 studies) | Not serious | Not serious | Serious^2^ | Not serious | Not serious | Moderate |
| Prognostic accuracy of hypocalcemia | 0.76 (0.53-0.90)  I^2^=96% | 0.53 (0.26-0.78)  I^2^=96% | 1,256 (6 studies) | Not serious | Serious^1^ | Serious^2^ | Not serious | Not serious | Low |
| Prognostic accuracy of hypokalemia | 0.25 (0.13-0.41)  I^2^=49% | 0.74 (0.59-0.85)  I^2^=84% | 684 (4 studies) | Not serious | Not serious | Serious^2^ | Not serious | Not serious | Moderate |

Quality of evidence for cross-sectional and cohort studies is graded starting at high quality for accuracy studies and downgraded or upgraded based on the following criteria: risk of bias, imprecision, inconsistency, indirectness and publication bias. 1. Downgraded by one level due to wide confidence intervals. 2. Downgraded by one level due to significant heterogeneity detected.
